# Impact of a pilot mHealth intervention on treatment outcomes of TB patients seeking care in the private sector using Propensity Scores Matching – Evidence collated from New Delhi, India

**DOI:** 10.1101/2023.12.05.23299517

**Authors:** Ridhima Sodhi, Vindhya Vatsyayan, Vikas Panibatla, Khasim Sayyad, Jason Williams, Theresa Pattery, Arnab Pal

## Abstract

Mobile health applications called Digital Adherence Technologies (DATs) are increasingly used for improving treatment adherence among Tuberculosis patients to attain cure, and/or other chronic diseases requiring long term and complex medication regimens. These DATs are found to be useful in resource limited settings because of their cost efficiency in reaching out to vulnerable groups (providing pill & clinic visit reminders, health information and awareness on the disease along with motivational messages and support to be retained in care) or those staying in remote or rural areas. Despite their growing ubiquity, there is very limited evidence on how they improve healthcare outcomes. We analyze the uptake of such an intervention in an urban setting (DS-DOST, powered by Connect for Life^TM^, Johnson & Johnson) among different patient groups accessing TB services in New Delhi, India, and subsequently assess its impact in improving patient engagement and treatment outcomes. This study aims to understand the uptake patterns of a digital adherence technology and its impact in improving follow ups and treatment outcomes among TB patients. Propensity choice modelling was used to create balanced treated and untreated patient datasets, before applying simple ordinary least square and logistic regression methods to estimate the causal impact of the intervention on the number of follow ups made with the patient and treatment outcomes.

After controlling for potential confounders, it is found that patients who installed and utilized DS-DOST application received an average of 6.4 (95% C.I. [5.32 to 7.557]) additional follow-ups, relative to those who did not utilize the application. This translates to a 58% increase. They also had 245% higher likelihood of a treatment success (Odds ratio: 3.458; 95% C.I. [1.709 to 6.996]). Descriptive results indicate that young females, and those suffering from pulmonary tuberculosis have a slightly higher propensity to use the CfL™ app, and benefit through their treatment duration.

**Author Summary:** The research tries to understand the impact of using cost-effective digital adherence tools, in improving treatment outcomes among patients diagnosed with drug-sensitive Tuberculosis (TB). As the treatment duration for TB is fairly long (at least 6 months) and complicated (multiple drugs, typically given in two distinct phases), there are challenges associated with ensuring treatment adherence. The research finds that digital tools such as a mobile application – can be a useful aid, albeit only when they are used in conjunction with the support of a healthcare worker. The digital tool analyzed, while sending medication reminders to patients, also enabled healthcare workers in tracking adherence for their assigned patients. The latter, as the research finds, ensured that they follow up with their patients to ensure adherence, resulting in increased odds of their getting a favourable treatment outcome. Further, the study underscores that a digital intervention used in isolation might not draw a favourable impact among patients – highlighting the role of healthcare workers and tailored interventions. In conclusion, digital adherence technologies can act as cost-effective measures in empowering healthcare workers to support their patients, and subsequently improve treatment outcomes.

## Introduction/Background

Digital adherence technologies (DATs) have increasingly evolved over time, and yet, evidence evaluating their impact on intended treatment or patient management outcomes is limited [1]–[3]. A previous study outlines multiple approaches of evidence generation for evaluating the efficacy of a mHealth solution, while also highlighting the inherent challenges associated [4]. One critical challenge highlighted here is the rapid pace of development of technologies, which could potentially entail the developers to improve or modify the intervention, thus making it difficult to accurately assess the impact of the mHealth intervention. Of the multiple approaches suggested to evaluate interventions, RCTs or randomized controlled evaluations were deemed to be the most common but far too long [5-7 years] in typical scenarios [4]. Another recommended approach called CEEBIT (Continuous Evaluation of Evolving Behavioural Interventions) discussed assessing multiple technologies, while continually removing inferior technologies from the competitive race [4], [5].

Notwithstanding the challenges, multiple studies have emphasized a need for evaluating DATs, and a simultaneous lack of the same [2], [6]–[8]. A systematic evaluation evaluating the role of mobile health interventions in enhancing interventions PPM (public-private mix) for TB care illustrated the increasing universe of such solutions [9]. However, none of the studies were found to evaluate the precise impact of such technologies on patient management (encompassing one or all aspects of pill reminders, patient follow ups, monitoring adherence, empowering patients with messages on disease awareness, side effects and health information) along with treatment outcomes. Even in more general studies documenting the usage of such technologies in TB care, there is a lack of quantitative evidence detailing the precise impact of such technologies on improving disease management or behaviour modification [1], [2].

Our study is aimed at closing this gap, by assessing the impact of a pilot intervention with Connect for Life™(CfL), a mobile-based digital adherence technology on patient management and treatment outcomes. The study utilizes propensity choice modelling to balance the test and control group, thus enabling precise estimates of the impact on treatment outcomes. The natural limitation of this method is that the test and control groups can only be balanced for the covariates on which the data is available. However, the magnitude and significance of results obtained, across different model specifications, render confidence to the inferences gathered.

## Methods

### Ethical Approval

CHAI’s Scientific and Ethical Review Committee (SERC) waived informed consent as anonymized programmatic data was utilized for the study. Additionally, all the individuals who enrolled in the CfL pilot did give an informed consent prior to enrolment in the pilot, through a written statement, and also gave consent via IVRS. In case of patients who were below 18 years of age, parents/guardians were primary participants, who provided written consent.

### DS-TB care services under Project JEET

The Joint Effort for Elimination of Tuberculosis (Project JEET) began in 2018 and is a large-scale private health sector engagement initiative for TB [10]. The services offered through the program are intended to reduce challenges which limit the Indian healthcare system in arresting TB transmission, facilitating access to appropriate TB care, and supporting TB patients throughout their treatment. As part of JEET, treatment coordinators facilitate the notification of newly diagnosed TB patients in a digital government tracking system called Nikshay, by liaising with private providers. This notification helps in tracking diagnosed patients and offering them a package of services provided by the National Tuberculosis Elimination program (NTEP). The patients also get regular counselling support by a designated treatment coordinator by way of in-person and telephonic follow ups, along with quality assured diagnostic services (Xpert Testing) and access to free government sponsored FDC (Fixed Dose Combination) drugs. The services are provided in close coordination with the treating physician, and as deemed appropriate by them. The program helps in limiting the onward transmission of disease through the combination of support described. At the time of initiation of the CfL pilot (November 2019), approximately 900 private providers in New Delhi were engaged with Project JEET as part of the Patient Provider Support Agency (PPSA) managed by the William J. Clinton Foundation (WJCF).

### Pilot Set up

The pilot was initiated in the month of November 2019, in three private care facilities in New Delhi, India, namely, Vinod Karhana Hospital, Ganga Ram Hospital, and St. Stephens hospital. The patients under this intervention were notified through Project JEET. The intervention was done in collaboration between William J Clinton Foundation (WJCF), TB Alert, India & Johnson & Johnson (J&J), wherein the latter developed and customized Connect for Life^TM^, a mobile application built to help treatment providers and healthcare workers in patient management. The digital intervention was voluntary in nature, wherein newly diagnosed TB patients (or/and their caregivers) were informed about the CfL application and were asked for consent to being a part of this program. New patients were enrolled in the pilot between 26^th^ November 2019 and 15^th^ March 2020. Patients who consented were provided with digital support offered by the CfL mobile application and a designated treatment coordinator, in addition to the standard services provided to patients managed under Project JEET. Two key elements of the pilot are described below:

1. **Connect for Life™**: This is a mobile, feature or smartphone-based health application system which utilizes a combination of IVRS (Interactive Voice Response System) and SMS (short messaging service) to help patients in remembering to take their medications, provide reminders for visiting the clinic for planned check-ups and medication refills, while also giving them health tips covering topics such as nutrition, significance of adherence, stigma, and community transmission. As of 2021, CfL is an Open-Source Platform [11] and can be downloaded and used by any organization or country. Some key features of the mHealth application include:

a. Facility to enrol both the patient and the caregiver
b. The user could modify the frequency of the pill/health-tip reminders between daily/weekly/monthly, and could also set up a preferred time during the day for receiving reminders
c. The adherence captured through the application were pulled in a dashboard, which could then be viewed by the treatment coordinator. This allowed the treatment coordinator to monitor patient’s adherence and follow up as needed
d. Option to opt out of service at any time during the treatment
2. **Treatment Coordinators**: Three healthcare workers were engaged for the pilot, each one responsible for patient management of the enrolled patients in one of the three private facilities. While all patients under Project JEET are assigned a treatment coordinator, these three treatment coordinators were also given training on how to use the mobile application and explain its functions and utilities to the patients/caregivers. Once patients consented to participate in the pilot, the treatment coordinator helped patients by activating their mobile telephone number with a unique password and explained how to use the phone and/or text messaging functionalities. A key differentiator of these treatment coordinators was that they had access to the CfL visualization dashboard, which enabled them to monitor the self-recorded medication adherence by patients using the application. This helped them in providing differentiated counselling to the patients, by optimizing the counselling and follow-ups based on the analytics provided by the dashboard. This potentially helped reduce the burden of patient management for the treatment coordinators. **Training of the treatment coordinators**: The three treatment coordinators engaged for the pilot were trained by the WJCF staff. They worked very similar to the treatment coordinators working under Project JEET, following the same counselling guides and protocols. Like other treatment coordinators (which are not employed specifically for this intervention), they were also responsible for updating patient’s demographic and treatment information in Nikshay.

### Study design

The quasi-experimental study compared the follow up regimen and treatment outcomes of patients who were part of the CfL pilot (test dataset), with patients who were not part of the pilot, but were notified during the same time period in the same three facilities (control dataset). Both test & control datasets were matched using propensity scores for ensuring robust measures.

### Data source(s)

There are two data sources utilized for this study. The first is data obtained from the CfL application, which consisted of information on 476 patients who were signed up for the pilot. The other source is programmatic data collected as part of the services rendered under Project JEET. The data collected as part of regular JEET operations included 1) TB patients demographic characteristics such as age, sex, and diagnosing district, 2) TB diagnostic and treatment information, including type of diagnostic test performed, pulmonary or extrapulmonary diagnosis, whether free drugs were provided, patients’ treatment outcome, and 3) number of follow-up contacts made by treatment coordinators. The data collected by the pilot intervention included information on utilization of the CfL application. This consisted of multiple metrics such as the preferable time for health tips delivery for each patient, whether or not they recorded the adherence, and the number of times they interacted with the IVRS for various services. It also had information on which of the 476 patients provided consent (or not) and if a patient stayed engaged with the CfL pilot through the course of their treatment.

### Data Selection (Inclusion & Exclusion criteria)

We considered data for patients that were enrolled in one of the three facilities where the pilot was conducted and were diagnosed of drug sensitive TB between 1^st^ October 2019 and 31^st^ March 2020. This also represents patients who were diagnosed before the COVID-19 pandemic started impacting health services operations in India (the first nation-wide lockdown in India was implemented on 24^th^ of March 2020). Among these, only those patients were considered who had had a treatment outcome assigned to them at the time this study was conducted. The selection criteria are described in more detail in Table 1 & 2. A total of 989 patients were enrolled, of which 276 enrolled in the CfL application, provided consent and utilized it through their treatment.

**Table 1.** CfL Test dataset: Selection criteria pathway.

| Reason for inclusion/exclusion | #Patients | Patients |
| --- | --- | --- |
|  | Excluded | retained |
| Patients enrolled | (+) 476 | 476 |
| Consent required | (-) 138 | 338 |
| Patients opted out of the application after using | (-) 18 | 320 |
| Patient died before treatment initiation | (-) 1 | 319 |
| Patients transferred/migrated to a different facility | (-) 35 | 284 |
| Treatment outcome is pending | (-) 7 | 277 |
| Treatment outcome should be one of cured, complete, death or treatment failure | (-) 1 | 276 |

**Table 2.**
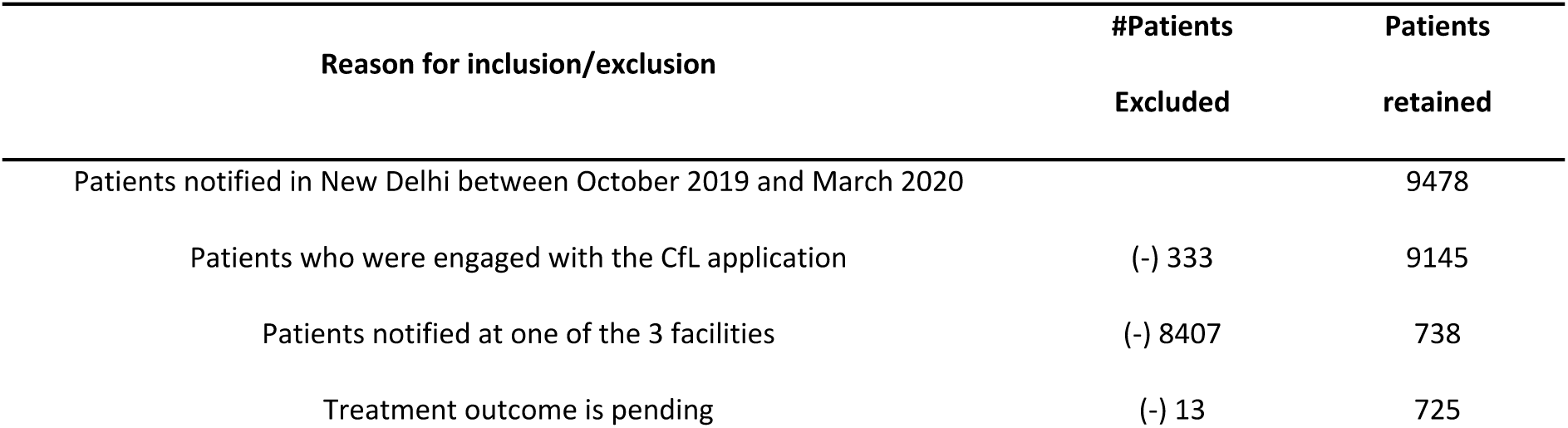
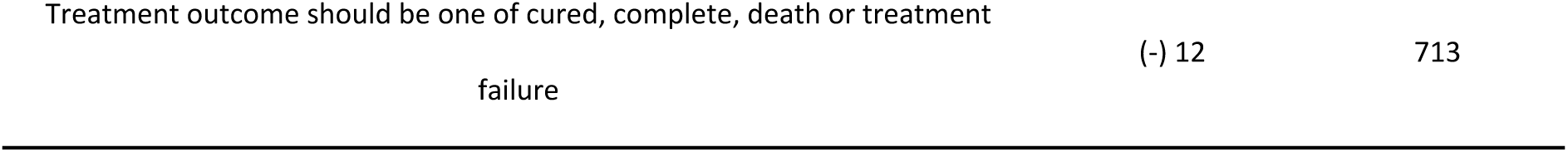
Control dataset: Selection criteria pathway.

### Model Theory

We fit multiple statistical models to understand how the CfL engagement impacted patients and their treatment outcomes. Our findings suggest that the combination of features provided by the CfL application contributed to the patient being more engaged with their treatment. Routine health tips, along with medication reminders, customized to be received at a chosen time of patient’s preference helped prevent challenges to continued treatment adherence, and encouraged the patient to stay connected with their treatment coordinator. Simultaneously, treatment coordinators utilized the platform to monitor patients’ adherence, and increased follow ups if and when the patients were found to lag behind in medication adherence. These factors then contribute to better treatment adherence, leading to better treatment outcomes (Figure 1). The study attempts to develop a precise estimate of the treatment effect of CfL on patients’ follow-ups and treatment outcome by using a combination of regression methods on a matched dataset built through propensity choice modelling.

**Figure 1:**
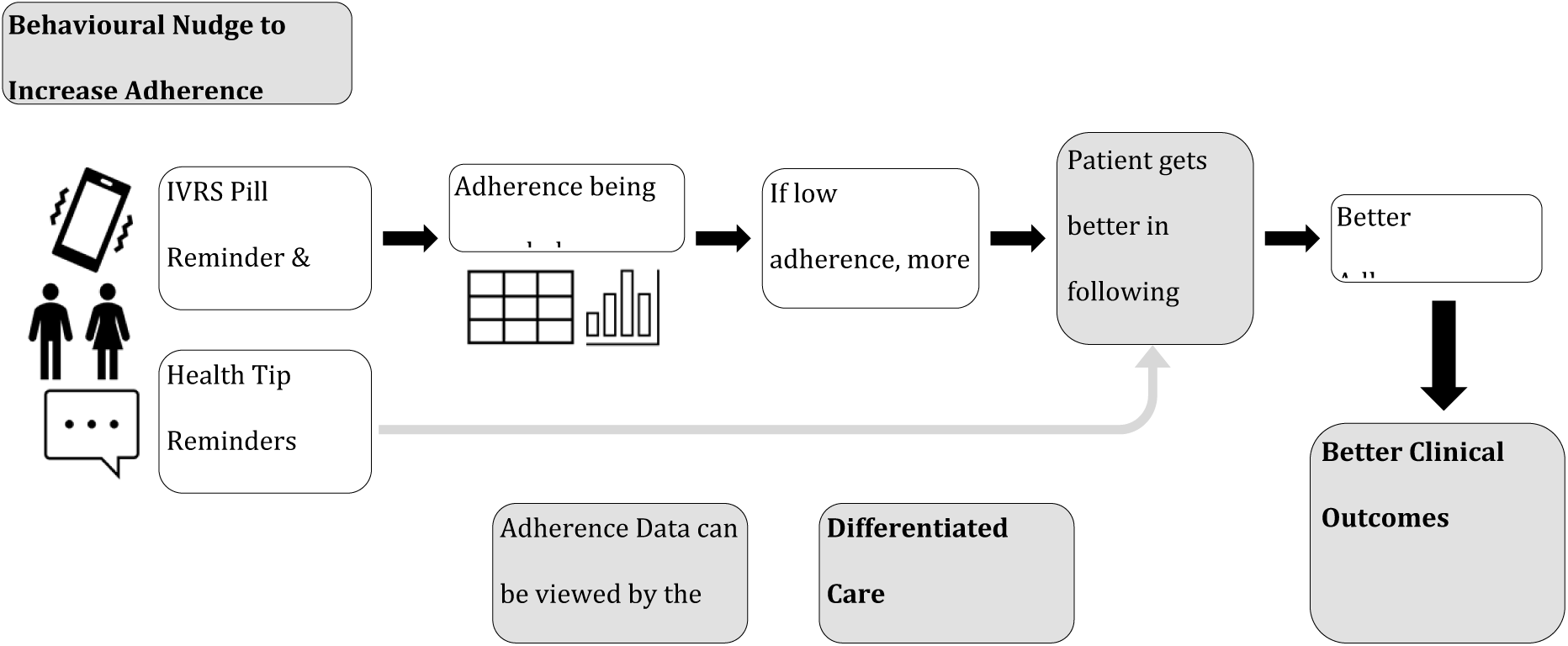
CfL Patient Care Cascade.

### Outcomes of Interest

The study has two primary outcomes, 1) patient follow-ups and 2) treatment outcomes. Patient follow ups refers to the number of times a patient spoke to or met with a treatment coordinator, and acts as a proxy for patient management. Treatment outcomes are analyzed as a binary variable in the study. Five outcomes are considered 1) treatment complete, 2) cured, 3) treatment failure, 4) death, or 5) lost to follow-up. The first two correspond to a successful outcome, and the last three correspond to an unsuccessful one. These outcomes are further described with clinical definitions in Appendix 1 [12].

### Propensity Choice Modelling

Effectively, all diagnosed TB patients who visited the three facilities during the pilot initiation were offered to enrol for the CfL intervention, meaning the data collected as part of this study was not randomized. This makes it difficult to access the average treatment effect (ATE) of the intervention on the outcomes of interest. While randomized experiments are a preferred choice to understand the causal effect of a treatment, running such an experiment is often cost intensive and consists of multiple ethical issues (primarily with respect to who receives the intervention), especially in studies concerning welfare and healthcare treatment effects [13]. Several studies have acknowledged the usage of matching methods to infer causal insights from observational data, specifically in the field of health care assessment [14], [15]. It is documented that creating a dataset which is matched on choice attributes provides an opportunity to estimate the average effect of the treatment as if it were a randomized experiment, which means if the access to CfL was randomly assigned to individual patients [16].

We utilized propensity score modelling to create a matched dataset comprised of treated patients (enrolled in the CfL) and untreated patients (not enrolled), which also included data on potential confounders for each individual [14], [17]–[20]. The propensity score refers to the conditional probability of a patient being enrolled in the pilot, given the values of all potential confounder [18]. This score was estimated for each patient in the full analytical dataset. These scores were then used to create comparable groups of people who were part of the pilot engagement (treated) and those who did not ever engage (untreated). The scores were adequate predictors of whether or not a patient enrolled in the pilot, illustrated in Appendix 2 (Figure SF1, Table S2, S3). We identified pairs of observations that have very similar propensity scores, but that differed in their treatment status (CfL or not), and employed the full matching algorithm, first developed by Rosenbaum (1991) [21] and illustrated by Hansen (2004) [22]. It uses all available individuals in the data by grouping the individuals into a series of matched sets (subclasses), with each matched set containing at least 1 treated individual (who received the treatment of interest) and at least 1 comparison individual (who did not). Full matching forms these matched sets in an optimal way, such that treated individuals who have many comparison individuals who are similar (on the basis of the propensity score) will be grouped with many comparison individuals, whereas treated individuals with few similar comparison individuals will be grouped with relatively fewer comparison individuals. The method is thus more flexible than traditional k:1 matching, in which each treated individual is required to be matched with the same number of comparison individuals (k), regardless of whether each individual actually has k good matches [23]. To counter any bias, we adopted two measures. First, we employed a calliper width of 0.2 for the age and district variables using nearest neighbour matching, meaning the matched pairs were a maximum of 0.2 standard deviations away from each other, which is described as ideal by previous studies [24]. Second, we employed exact matching on four variables: 1) proportion of males, 2) proportion of extra pulmonary cases, 3) proportion of patients diagnosed using Xpert testing, and 4) access to free drugs.

### Statistical modelling

Using the matched dataset, we fit fixed-effects ordinary least squares (OLS) regression and fixed-effects logistic regression models to estimate the impact of CfL engagement on the number of follow-ups made with the patient and the likelihood of a successful treatment outcome, respectively. As covariates in the OLS regression model, we fitted a series of models, sequentially including CfL engagement, diagnosing facility, sex, age category (0 to 5, 6 to 15, 16 to 19, 20 to 45, 46 to 65, and ≥ 66 years), TB type (pulmonary or extra pulmonary), whether Xpert diagnostics were used, access to free drugs, and the quarter in which the diagnosis was made. The logistic regression model was fit to assess the likelihood of a patient receiving a successful outcome at the culmination of treatment. The same covariates went into the logistic regression model.

Diagnosing quarter was included in the model OLS and logistic regression models to control for seasonal program-related influences of patient care and adherence to treatment. Interaction effects on diagnosing facility and diagnosing quarter were considered in both models to account for the simultaneous effect of these two variables on the dependent variables [25].

To establish the linkage between follow ups and treatment outcomes, a logistic model was fit with follow ups as an additional dependent variable, while including for the status of CfL engagement and other covariates.

### Sensitivity Analysis

Multiple sensitivity analyses were conducted to understand the impact of the CfL engagement on follow ups and treatment outcomes. These are illustrated in the forest plots (Figures 6 and 7), tabular results of which are provided in Appendix 5. Our results were robust to specifications which excluded cases where treatment outcome resulted in lost to follow up, or when we ran facility specific models. The results were also robust to alternative matching methods, wherein we utilized nearest neighbor matching, along with exact matching for selected variables (Appendix 6). The latter resulted in 204 matched pairs.

### Statistical software

Analysis was conducted in R 2022.07.01. ‘MatchIt’ package [26], [27] was used for the propensity score matching procedure, ‘broom’, ‘cobalt’, and ‘gtsummary’ packages were used for visualizing fitted and residual values, generating balance plots from propensity choice modelling, and generating summary statistics, respectively. Packages used for data cleaning, preparing the analytical datasets, measuring skewness, and visualizing results were ‘dplyr’, ‘tidyr’, ‘moments’, and ‘ggplot2’.

## Results

Tables 3 provides the demographic and clinical profiles of patients in the analytical dataset (989 patients) and matched dataset (944 patients). Table 4 further provides this information, albeit segregated by the pilot engagement status. The matching process using propensity scores brought the standardized propensity score difference between the treated and control group from 0.28 to 0, while balancing the mean difference between other covariates (Appendix 3). Within the matched dataset, 56% patients were male, 20% of patients were diagnosed using Xpert testing, 56% patients had extra pulmonary TB, and 88% of patients had a successful treatment outcome recorded.

**Table 3.**
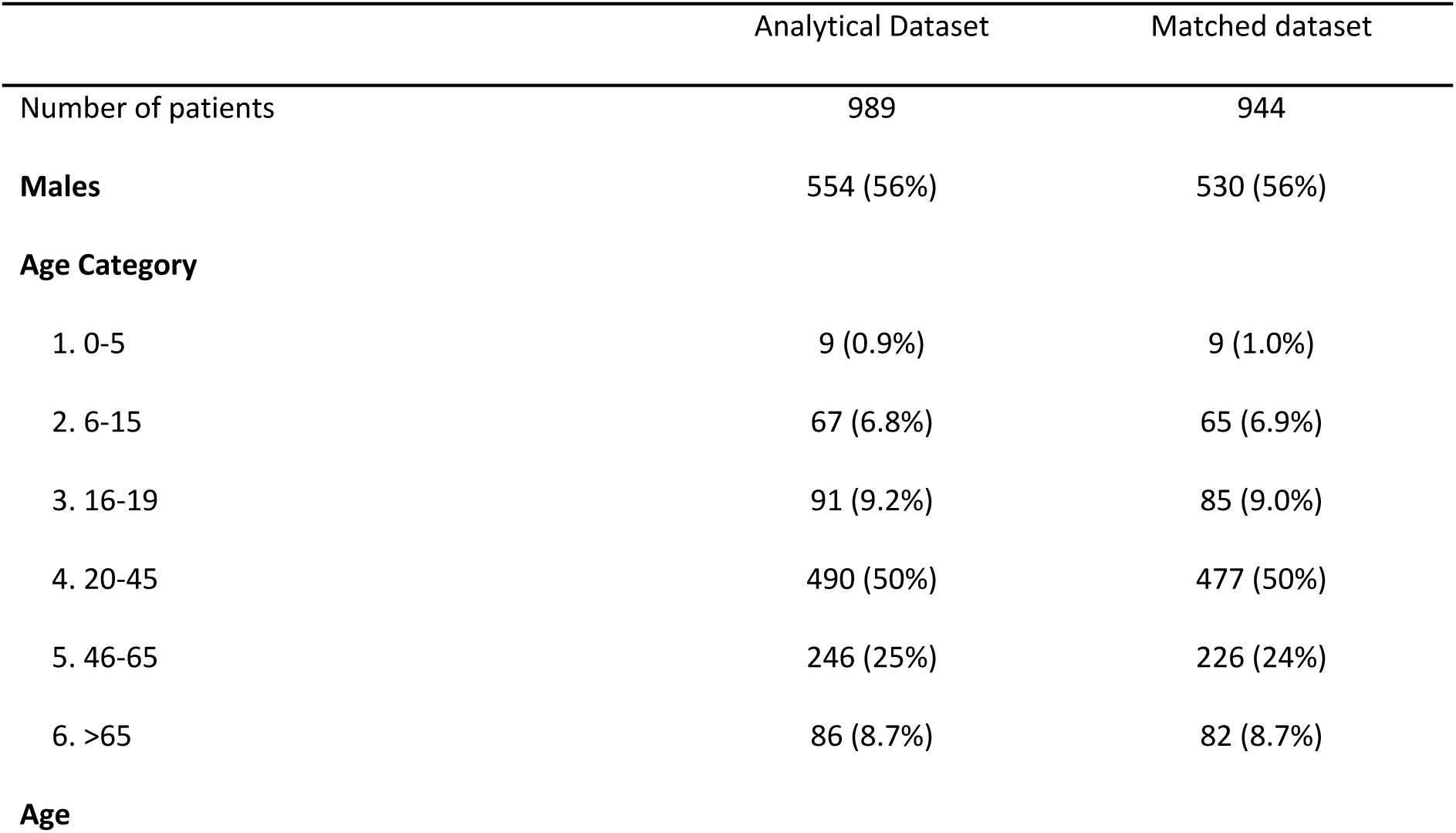
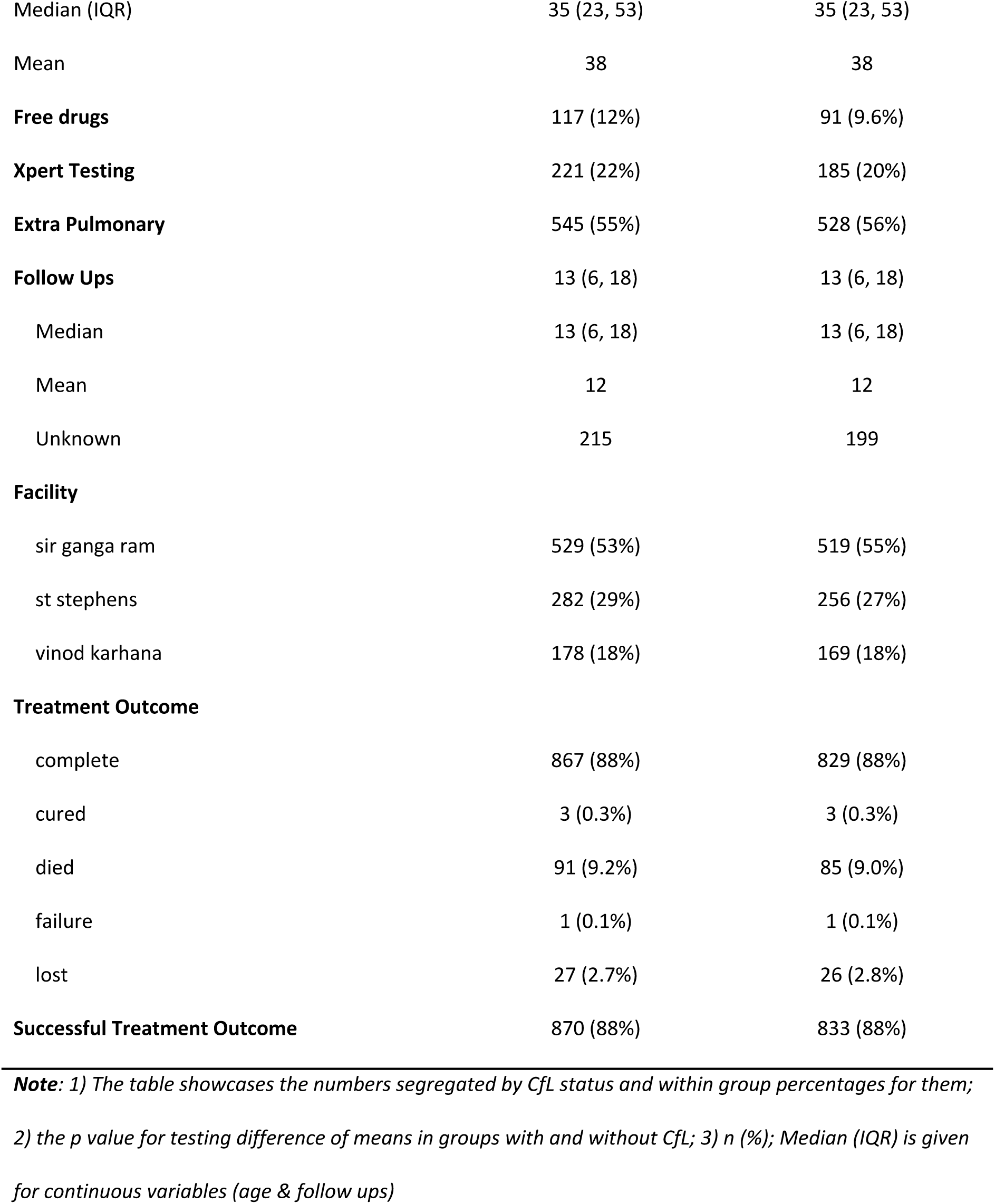
Summary statistics for dataset before and after matching.

|  | Analytical Dataset | Matched dataset |
| --- | --- | --- |
| Number of patients | 989 | 944 |
| **Males** | 554 (56%) | 530 (56%) |
| **Age Category** |  |  |
| 1. 0-5 | 9 (0.9%) | 9 (1.0%) |
| 2. 6-15 | 67 (6.8%) | 65 (6.9%) |
| 3. 16-19 | 91 (9.2%) | 85 (9.0%) |
| 4. 20-45 | 490 (50%) | 477 (50%) |
| 5. 46-65 | 246 (25%) | 226 (24%) |
| 6. >65 | 86 (8.7%) | 82 (8.7%) |
| **Age** |  |  |
| Median (IQR) | 35 (23, 53) | 35 (23, 53) |
| Mean | 38 | 38 |
| **Free drugs** | 117 (12%) | 91 (9.6%) |
| **Xpert Testing** | 221 (22%) | 185 (20%) |
| **Extra Pulmonary** | 545 (55%) | 528 (56%) |
| **Follow Ups** | 13 (6, 18) | 13 (6, 18) |
| Median | 13 (6, 18) | 13 (6, 18) |
| Mean | 12 | 12 |
| Unknown | 215 | 199 |
| **Facility** |  |  |
| sir ganga ram | 529 (53%) | 519 (55%) |
| st stephens | 282 (29%) | 256 (27%) |
| vinod karhana | 178 (18%) | 169 (18%) |
| **Treatment Outcome** |  |  |
| complete | 867 (88%) | 829 (88%) |
| cured | 3 (0.3%) | 3 (0.3%) |
| died | 91 (9.2%) | 85 (9.0%) |
| failure | 1 (0.1%) | 1 (0.1%) |
| lost | 27 (2.7%) | 26 (2.8%) |
| **Successful Treatment Outcome** | 870 (88%) | 833 (88%) |

**Note:** 1) The table showcases the numbers segregated by CfL status and within group percentages for them; 2) the p value for testing difference of means in groups with and without CfL; 3) n (%); Median (IQR) is given for continuous variables (age & follow ups)

**Table 4.** Summary statistics for dataset before and after matching; segregated by the CfL^TM^ pilot engagement status.

| Pilot engagement | Analytical Dataset |  |  | Matched dataset |  |  |
| --- | --- | --- | --- | --- | --- | --- |
|  | (N = 989) |  |  | (N = 944) |  |  |
|  | no CfL | CfL | p-value | no CfL | CfL | p-value |
| **Number of patients** | 713 | 276 |  | 694 | 250 |  |
| **Males** | 406 (57%) | 148 (54%) | 0.3 | 398 (57%) | 132 (53%) | 0.2 |
| **Age Category** |  |  |  |  |  |  |
| 1. 0-5 | 6 (0.8%) | 3 (1.1%) |  | 6 (0.9%) | 3 (1.2%) |  |
| 2. 6-15 | 45 (6.3%) | 22 (8.0%) |  | 45 (6.5%) | 20 (8.0%) |  |
| 3. 16-19 | 51 (7.2%) | 40 (14%) |  | 51 (7.3%) | 34 (14%) |  |
| 4. 20-45 | 336 (47%) | 154 (56%) |  | 334 (48%) | 143 (57%) |  |
| 5. 46-65 | 200 (28%) | 46 (17%) |  | 186 (27%) | 40 (16%) |  |
| 6. >65 | 75 (11%) | 11 (4.0%) |  | 72 (10%) | 10 (4.0%) |  |
| **Age** |  |  | <0.001 |  |  | <0.001 |
| Median (IQR) | 38 (24, 56) | 30 (20, 42) |  | 37 (24, 56) | 30 (20, 42) |  |
| Mean | 40 | 33 |  | 40 | 33 |  |
| **Free drugs** | 48 (6.7%) | 69 (25%) | <0.001 | 41 (5.9%) | 50 (20%) | <0.001 |
| **Xpert Testing** | 105 (15%) | 116 (42%) | <0.001 | 88 (13%) | 97 (39%) | <0.001 |
| **Extra Pulmonary** | 408 (57%) | 137 (50%) | 0.031 | 402 (58%) | 126 (50%) | 0.040 |
| **Follow Ups** |  |  |  |  |  | <0.001 |
| Unknown | 185 | 30 |  | 173 | 26 |  |
| Median (IQR) | 11 (4, 16) | 18 (13, 20) | <0.001 | 11 (4, 16) | 18 (13, 20) |  |
| Mean | 10 | 16 |  | 10 | 16 |  |
| **Facility** |  |  | <0.001 |  |  | <0.001 |
| sir ganga ram | 454 (64%) | 75 (27%) |  | 447 (64%) | 72 (29%) |  |
| st stephens | 153 (21%) | 129 (47%) |  | 143 (21%) | 113 (45%) |  |
| vinod karhana | 106 (15%) | 72 (26%) |  | 104 (15%) | 65 (26%) |  |
| **Successful treatment outcome** | 608(85%) | 262(95%) | <0.001 | 595 (86%) | 237 (95%) | <0.001 |

**Notes:** 1) The table showcases the numbers segregated by Cfl status, and within group percentages for them; 2) For binary/character variables, values represent the number of patients enrolled, and value in parentheses represents share or %; 3) For continuous values, the number represents the median, and the values in parentheses represents the Interquartile Range; 4) Pearson's Chi-squared test and Kruskal-Wallis rank sum test is conducted for p value

### Follow-ups with patients

Within the matched dataset, patients with the CfL^TM^ engagement received more follow-ups from treatment coordinators (Mean=16.3, Median (IQR): 18 (13,20)) than patients who were not engaged with the pilot (Mean = 10.3, Median (IQR): 11 (4, 16)). These are also illustrated in Figure 2 by way of a box plot distribution. We fit a series of five regression models that progressively added patient-level covariates, fixed-effects for facility, patient level covariates, fixed-effects for quarter of diagnosis, and an interaction between facility and quarter of diagnosis (Table 6). All five models revealed statistically significant effect of the pilot engagement on follow-ups, with Model E controlling for all available potential confounders and an interaction term on diagnosing facility and quarter. Here, an average treatment effect (ATE) in terms of 6.4 additional follow-ups (95% C.I. = 5.295 to 7.540) was found for patients enrolled in the pilot relative to those who were not. This is equivalent to a 62% increase in mean follow-ups (58% increase if we compare median values).

**Figure 2:**
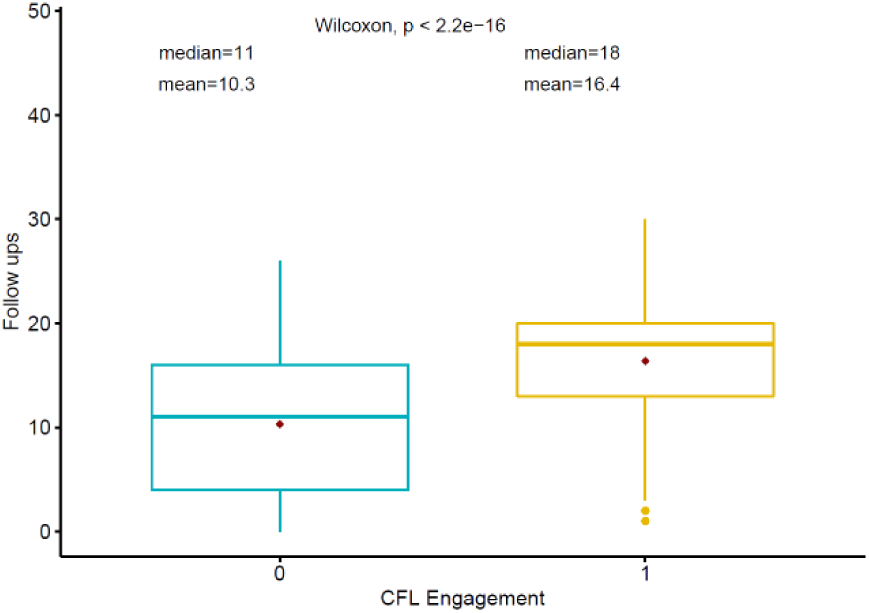
Box plot showing the difference in follow ups made with patients; based on whether they were a part of the CfL pilot; Matched Dataset; N = 946. Note: The diamond dot represents mean, and the box boundary represents the interquartile distribution

**Figure 3:**
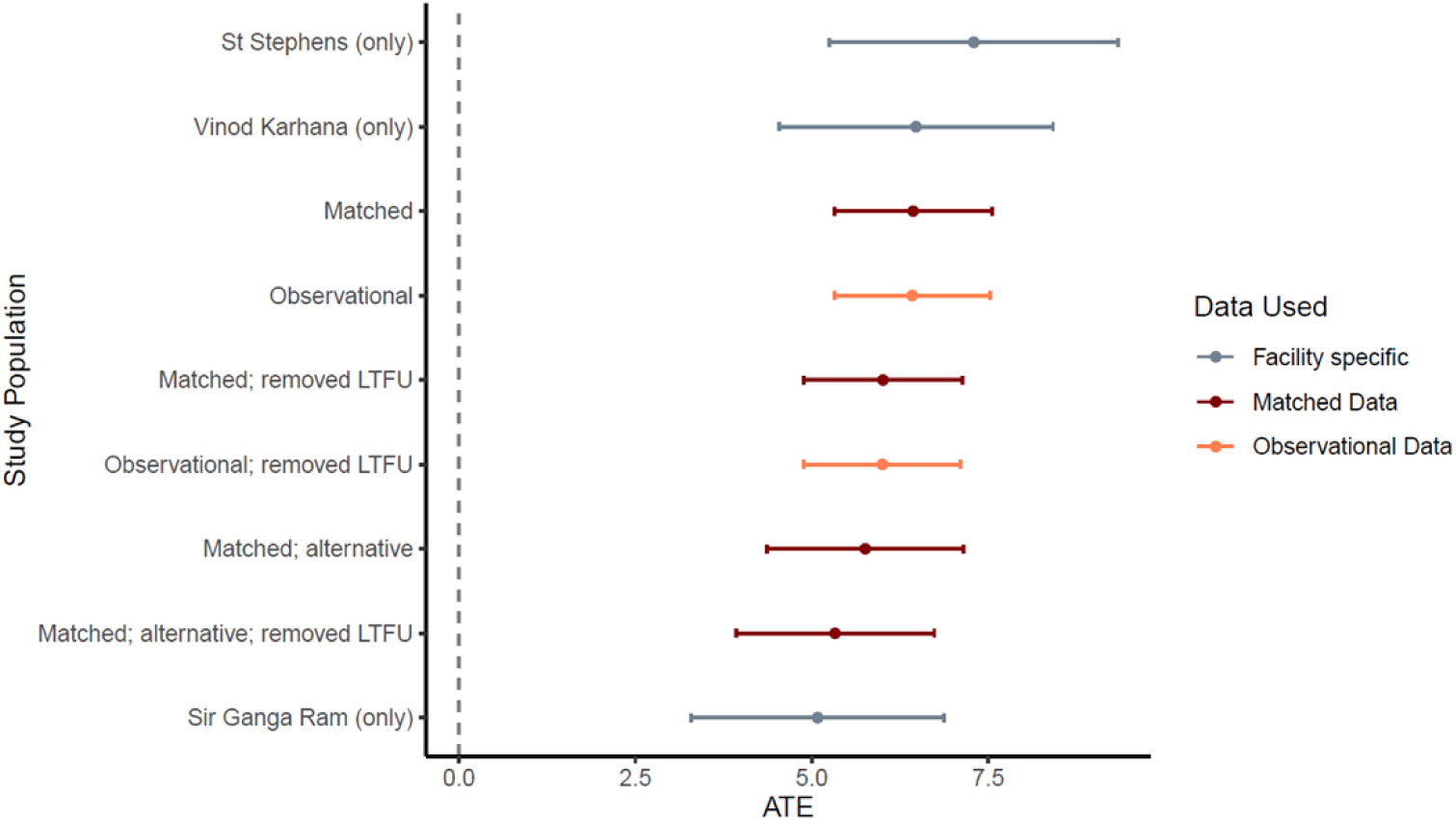
Forest Plot for OLS Regression; Impact of CfL on follow Ups Effect Size by sub population or Matching Specification. Note: 1) LTFU refers to Lost to follow up; 2) Dotted line at X = 0 helps in visualizing the sub populations which reveal a significant impact (or not) of CfL on follow ups outcomes; 3) All facility specific models are fitted on the matched dataset; 4) Matched (alternative) refers to an alternative matching specification (Appendix 7)

**Figure 4:**
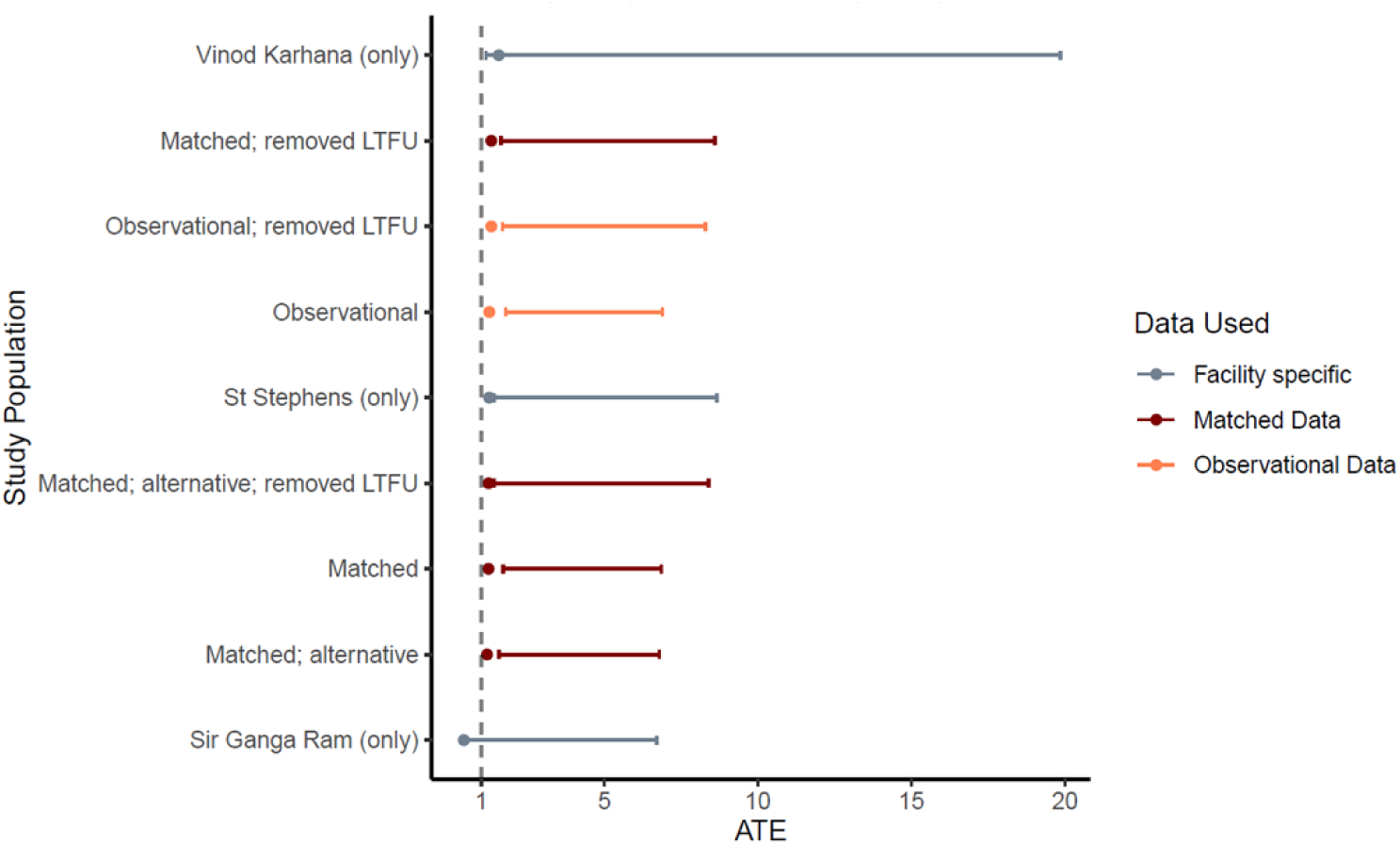
Forest Plot; Sensitivity Analysis Logistic Regression; Impact of CfL provision on Treatment Outcomes. Note: 1) OR Ratios are displayed along with 95% C.I.; 2) Dotted line at X = 1 illustrates the sub-populations which reveal a significant impact of CfL on treatment outcomes (to the right) or not (to the left) 3) LTFU refers to lost to follow-up; 4) All facility specific models are fitted on the matched dataset; 5) Matched (alternative) refers to an alternative matching specification (Appendix 7)

**Table 5.** OLS regression results showing impact of the pilot CfL engagement on number of follow-ups with patients using matched dataset; N = 745.

|  | Model A | Model B | Model C | Model D | Model E |
| --- | --- | --- | --- | --- | --- |
| CfL Engagement | 5.995 | 6.128 | 6.131 | 5.943 | 6.417 |
| 95% C.I. | (4.986, 7.003) | (5.070, 7.185) | (4.963, 7.298) | (4.768, 7.119) | (5.295, 7.540) |
| + Facility fixed effects |  | Yes | Yes | Yes | Yes |
| + Additional Covariates |  |  | Yes | Yes | Yes |
| + Diagnosing Quarter fixed effects |  |  |  | Yes | Yes |
| + Facility by Diagnosing quarter interaction |  |  |  |  | Yes |
| Observations | 745 | 745 | 745 | 745 | 745 |
| R<sup>2</sup> | 0.150 | 0.154 | 0.176 | 0.180 | 0.216 |
| Adjusted R<sup>2</sup> | 0.148 | 0.151 | 0.162 | 0.166 | 0.200 |
| Residual Std. Error | 6.561 (df = 743) | 6.552 (df = 741) | 6.507 (df = 732) | 6.494 (df = 731) | 6.359 (df = 729) |
| F Statistic | 130.748 (df = 1; 743) | 45.098 (df = 3; 741) | 13.026 (df = 12; 732) | 12.374 (df = 13; 731) | 13.417 (df = 15; 729) |
|  | (df = 1; 745) | (df = 3; 743) | (df = 12; 734) | (df = 13; 733) | (df = 15; 731) |

Note: a) 95% C.I. based on robust standard errors; b) All models were fitted on matched dataset; c) \*p<0.1;
\*\*p<0.05; \*\*\*p<0.01

### Treatment Outcomes

A series of fixed-effects logistic regression models (Table 7) revealed a statistically significant greater likelihood of a successful treatment outcome for patients who was enrolled in the CfL pilot, relative to those who were not. Model E includes controls for all available covariates and reveals 242% higher odds (OR = 3.415; 95% C.I. [1.701 to 6.857]) of a successful outcome for patients who was enrolled in the pilot relative to others.

**Table 6.**
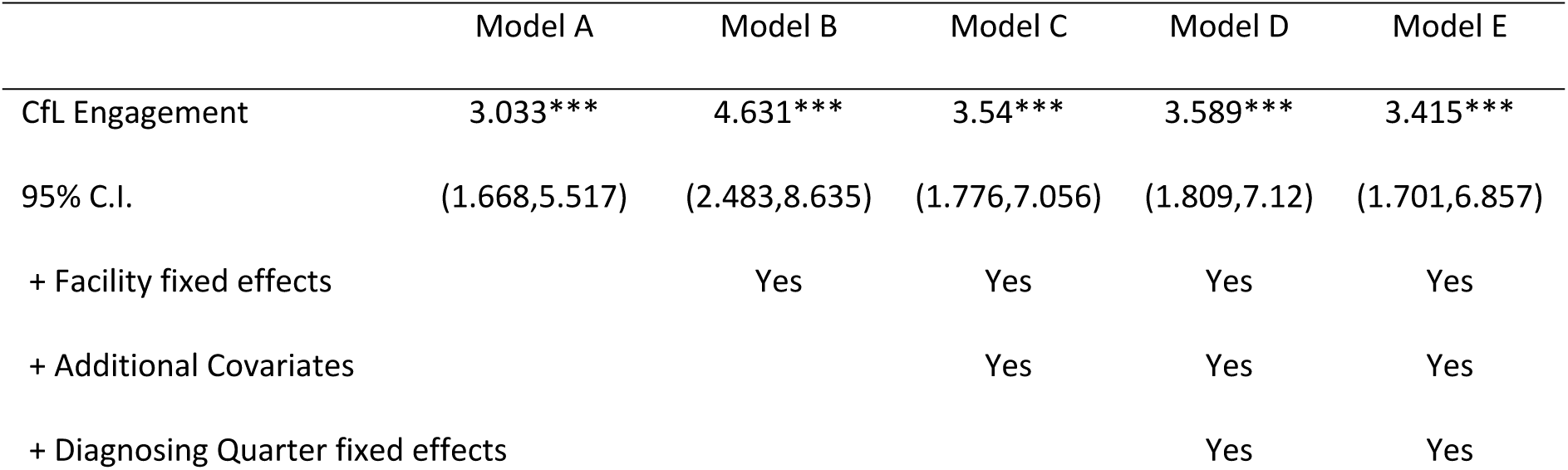
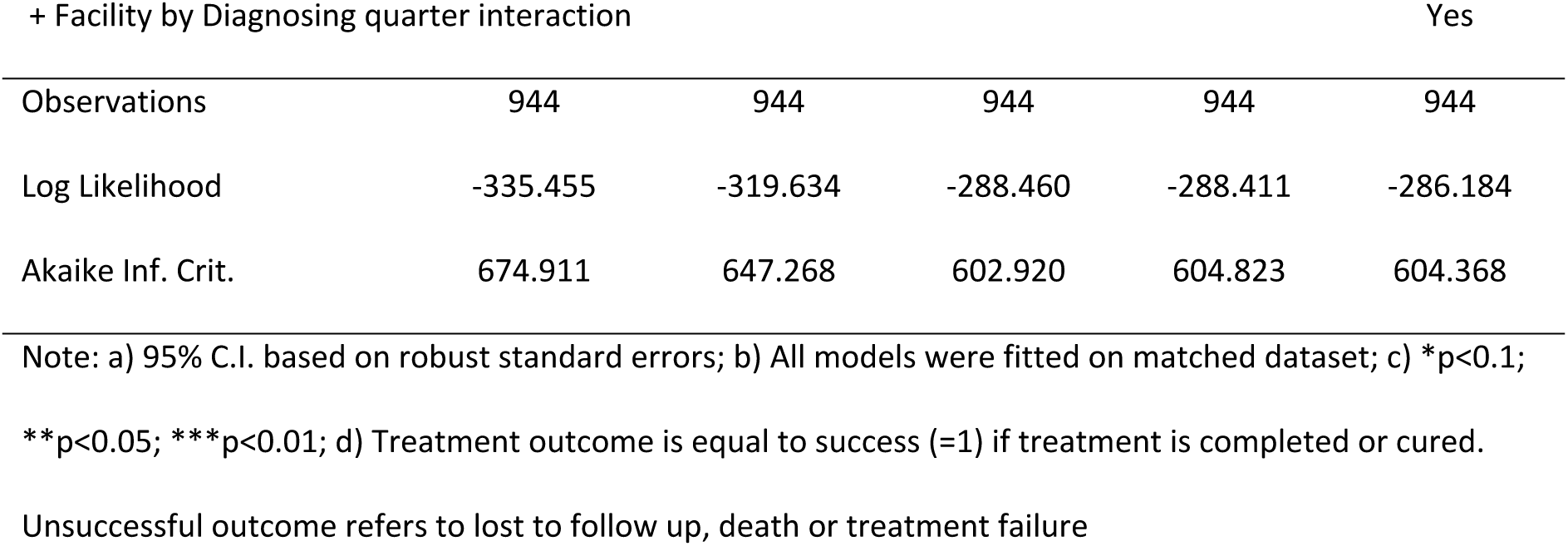
Logistic regression results showing impact of the pilot CfL engagement on treatment outcomes using matched dataset; N = 944.

### Link between CfL, follow ups, and treatment outcomes

Including follow ups as a covariate in the logistic model reduces the size and significance of the coefficient on CfL (Table 7.1). It also reveals a statistically significant coefficient on the follow ups, estimating a 24% increased likelihood of a successful outcome, for every unit increase in follow ups with the patient. Results from this specification (Table 7.1), along with the model revealing a significant impact of CfL drugs on follow ups (Table 6), lead us to conclude that the CfL engagement is leading to better treatment outcomes, primarily through their impact on the number of follow ups made with the patient.

**Table 6.1.**
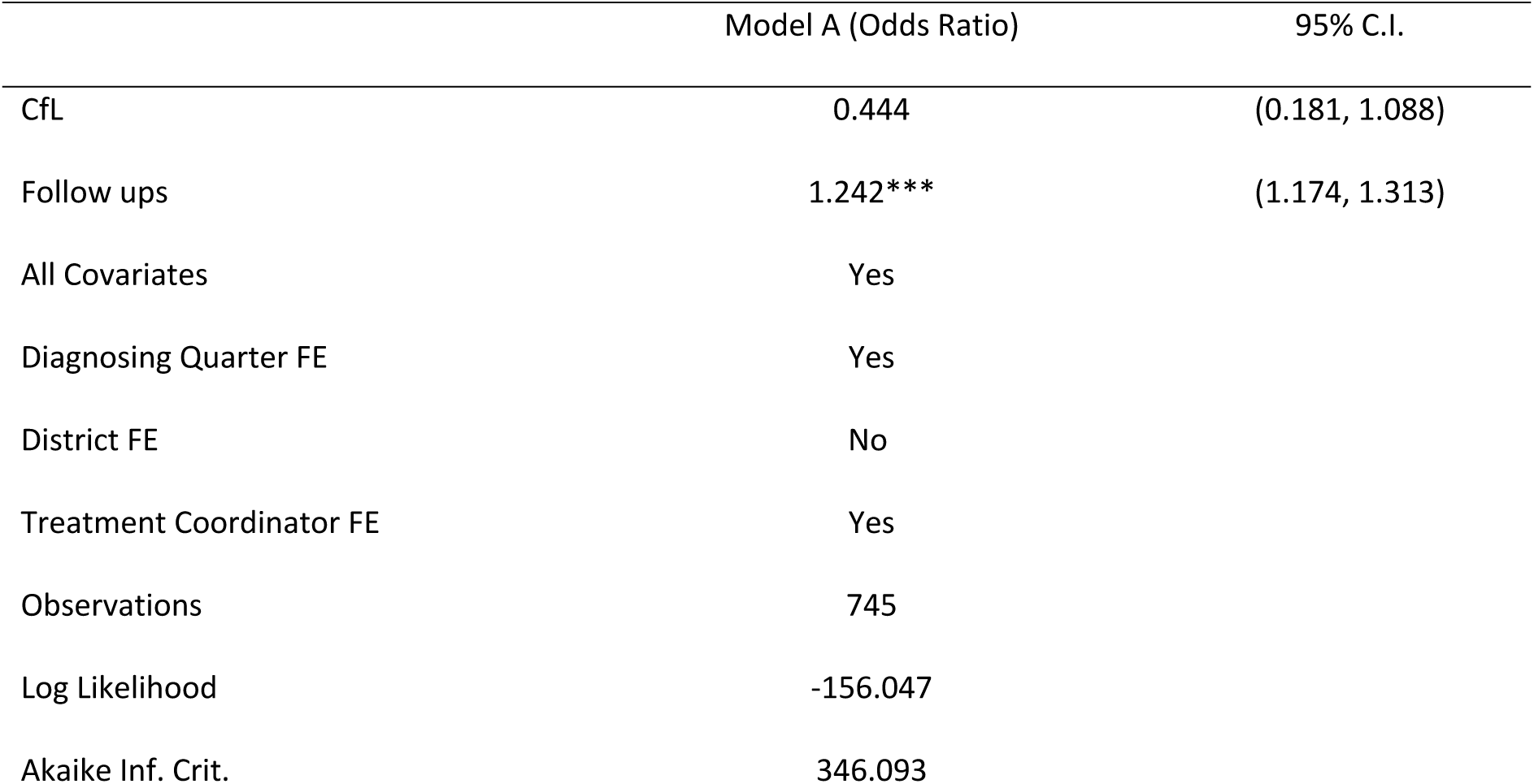
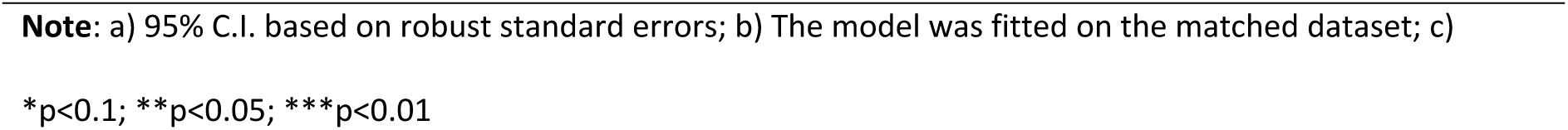
Logistic regression results showing impact of CfL on treatment outcomes using matched dataset; N = 745; including follow ups as a covariate.

|  | Model A (Odds Ratio) | 95% C.I. |
| --- | --- | --- |
| CfL | 0.444 | (0.181, 1.088) |
| Follow ups | 1.242*** | (1.174, 1.313) |
| All Covariates | Yes |  |
| Diagnosing Quarter FE | Yes |  |
| District FE | No |  |
| Treatment Coordinator FE | Yes |  |
| Observations | 745 |  |
| Log Likelihood | -156.047 |  |
| Akaike Inf. Crit. | 346.093 |  |

**Note:** a) 95% C.I. based on robust standard errors; b) The model was fitted on the matched dataset; c)
\* $p < 0.1$ ; \*\* $p < 0.05$ ; \*\*\* $p < 0.01$

## Discussion

To our knowledge, this is the first study assessing the impact of a DAT intervention on TB patients seeking care in the Indian private sector using a quasi-experimental approach. Our findings illustrate that patients who used the CfL application had a significantly higher likelihood of completing their treatment successfully, likely caused by an increase in follow ups associated with the usage of the application. The results stay robust after employing robust propensity score matching methods and a series of sensitivity analysis.

While digital adherence technologies (DATs) have been increasingly used for improving patient behaviours across the globe, their particular usage in India has remained underutilized, in part due to a lack of proper regulation and implementation [28]. Additionally, in-person or telephonic follow ups, though a gold standard to improve all aspects of patient management, can be burdensome for developing countries such as India because of high patient load, stagnant workforce and high number of patients living in rural and remote areas [29]. A study in Uganda illustrated the role played by such DATs where face to face counselling and social support is expensive because of a lack of financial resources and difficulties in transportation [30]. Wider usage of digital applications has the potential to mitigate these challenges [3]. Studies have highlighted the role of effective communication through mHealth technologies in India, which could bridge the gap between patient and medical staff interaction [31]. Treatment for tuberculosis is particularly complex and long drawn, which further intensifies the need for different novel methods to ensure patients adhere to the treatment through different challenges[32]. A modelling analysis estimates that such DATs, if employed in the public sector alone, can potentially reduce TB incidence by 7.3% over a course of 10 years, and by 16% if also deployed in the private sector, albeit under idealized settings [33]. Another study conducted in Bengaluru, India highlighted the role of an mHealth application (Kill TB) in using reminders to improve patient adherence [34].

Earlier examples of digital interventions to manage adherence include 99DOTS [35], Video Directly Observed therapy (VDOT) or Video Observed therapy (VOT) and Event Monitoring device for medication support (EMM) [36]. The current landscape is still an evolving one, and evolving iterations of these devices and solutions are being piloted for understanding their use cases. Recent evaluations of the Medical Event Reminder Monitor (MERM) box [37] and TMEAD [38], both applications of EMM, has found favourable outcomes among patients using these solutions, albeit stating challenges with respect to the actual usability [37]–[41].

Previous studies have recommended these DATs to be used to support a larger patient-management system, wherein differentiated counselling or switching to DOT can be possible options in case of non-adherence [42]– [44]. While all of these interventions included a dashboard solution to enable patient monitoring by healthcare workers, most evaluative studies have not specifically evaluated the impact thereof [2], [45]. In our study, we find that patients enrolled in the CfL intervention received a higher number of follow ups. Earlier studies have highlighted the role of enabling differentiated care [46] and the importance of human-interactions in improving success from DATs [47]–[49]. A previous systematic review talks about using such DATs to enable differentiated care, and more intensive face-to-face engagement as and when required [3]. With a significant amount of mixed evidence around the impact of DATs, results from this analysis suggests that digital technologies might show little impact if used in isolation. This follows from the fact that medication non-adherence is a complex issue with multiple contributing factors [50], [51], and a tailored intervention is required to draw positive impact from any singular technological solution [3].

## Conclusions

The results from this analysis are significant in illustrating the impact of the CfL solution in improving the ability of healthcare workers to counsel patients effectively, while simultaneously improving a patient’s engagement with their treatment by providing a combination of services (health tips delivery, drug medication reminders, clinic visit reminders). Digital interventions such as these serve as a low-cost method to improve patient behaviours with respect to continuing treatment. They help reach out to population groups who do not have easy access to a clinic, and may be living at far-off remote, rural, or hard to reach areas which make transportation costs a huge barrier in accessing clinician services. They also help reduce stigma and generate awareness among patients and caregivers, potentially improving patient attitudes to treatment and care. Similar interventions, which help optimize patient care, could also have the potential to reduce the psychological burden borne by healthcare workers in resource-constrained settings. Future scaled deployments of such technologies need to consider the importance of a multi-faceted intervention, combining the elements of technology and human-centred approach in order to improve treatment outcomes for patients.

## Limitations

Our analysis, while strongly suggesting the impact of the CfL intervention, has several limitations which warrant further research. **First**, our analysis does not investigate the specific implementation challenges witnessed by the program team, which would be essential to initiating a scale up of the same across a wider geography or/and a greater number of facilities. Some of the impediments noted by the program staff included a) disruptions in internet/telephone-network at the home location of patients, and b) patients complaining about redundant or repetitive content in health tip deliveries, and c) lengthy process to record their adherence into the system. It is worth noting that broadly, the CfL application was flexible to individual patients’ needs. For instance, a patient or caregiver could modify the frequency of their reminders, set up a preferred time for the same, while also selecting the topics on which they would require nudges (pill reminders, doctor visit reminders, adherence report, nutrition, and plausible side effects, among other things.). Nevertheless, quickly responding to similar challenges within a built-in software solution can be cumbersome, especially if deployed across a larger patient group. Notwithstanding the plausible challenges, several of the commonly found difficulties with using such solutions (application crash, data recording errors, erroneous or harmful information in terms of health tips) [52] were not reported from patients or the program staff using this application. **Second**, while the results are accompanied by a facility-level sensitivity analysis, our research does not attempt to find reasons for heterogeneity in results obtained or delve into the specifics of implementation in these different facilities. It is worth noting that Vinod Karhana is a relatively smaller facility, with respect to the number of patients catered to, when compared with Sir Ganga Ram hospital and St. Stephens hospital. Both Sir Ganga Ram and St. Stephens are situated in central Delhi, making it relatively easier for patients to access them by various forms of public transportation. A more detailed, and perhaps qualitative narrative of how such interventions might impact patients visiting such diverse facilities is warranted. The same can help inform differential enrolment and investment strategies, which can be more efficient in utilizing such DATs for TB, as well as other diseases involving long and complicated treatment regimens. **Third**, while we investigate the impact of the intervention on treatment outcomes, we do not compare the adherence recorded among patients, where the latter is the primary indicator being measured by the CfL application. Our reservation against measuring the adherence stems from the fact that it is self-reported by the patient and is likely to have some bias. Our analysis adopted a more outcomes-based approach, by directly evaluating the impact on the number of follow ups made, and the likelihood of successful treatment completions. However, research assessing medication adherence could be worthwhile in understanding more specific barriers to patient engagement and the successful deployment of such interventions. **Fourth**, we use a derived dichotomous outcome variable to understand the impact of the intervention on treatment outcomes. Here, unsuccessful outcomes include treatment failure, death, and lost to follow-up, and each of these outcomes may have their own risk profiles. All patients who had a treatment interruption greater than one month in duration are considered as being lost to follow-up. However, it cannot be determined if patients continued the treatment later, and if so, whether they were able to complete the treatment with a positive outcome. Hence, including lost to follow-up has the potential to bias these results. Some previous studies have not included lost to follow-up in their analyses for similar reasons [53]. However, we remain conservative and followed the baseline criteria of including patients who were under the active management of a treatment coordinator, and had their outcomes reported at least a month after the date of diagnosis. Additionally, in our particular analysis, lost to follow-up makes up 2.7% and 2.8% of our analytical and matched datasets, respectively. Including lost to follow-up in analysis where these cases make less than <5% of the overall population generally leads to little bias [54]. Sensitivity analysis excluding patients with lost to follow up as a treatment outcome (Appendix 6) supported the primary findings with high statistical significance. Regardless, further research is warranted to fully understand the differential risk profile of private sector TB patients, including the drivers of lost to follow-up and treatment failure. **Lastly**, majority of patients reported treatment completion which was based on the provider declaring that patient need not take any more medications. Since cure rates are low due to lack of smear testing in the private sector, the metric of successful treatment completion itself has certain limitations.

## Data Availability

The data utilized for the work is available in a Github repository: https://github.com/ridhimasodhi/CfL-Study

## List of abbreviations

DAT: Digital Adherence Technology
ATE: Average treatment effect
CI: Confidence interval
FDC: Fixed dose combination
IQR: Interquartile range
JEET: Joint Effort for Elimination of Tuberculosis
OLS: Ordinary least squares
PPSA: Private Provider Support Agency
TB: Tuberculosis
NTEP: National Tuberculosis Elimination Program
NSP: National Strategic Plan for Elimination of Tuberculosis

## Appendix 1 Definition of treatment outcomes

**Table S1.**
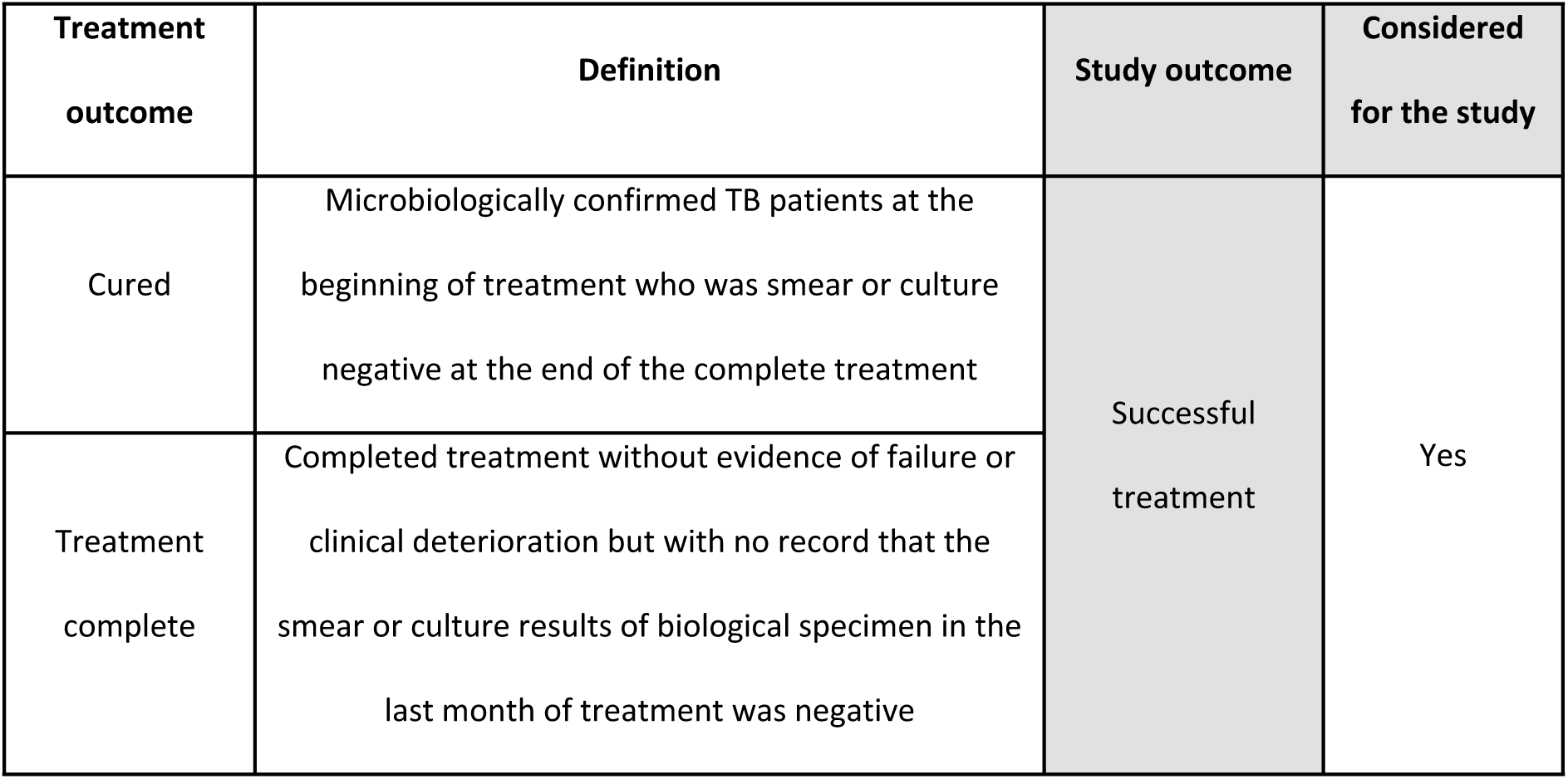
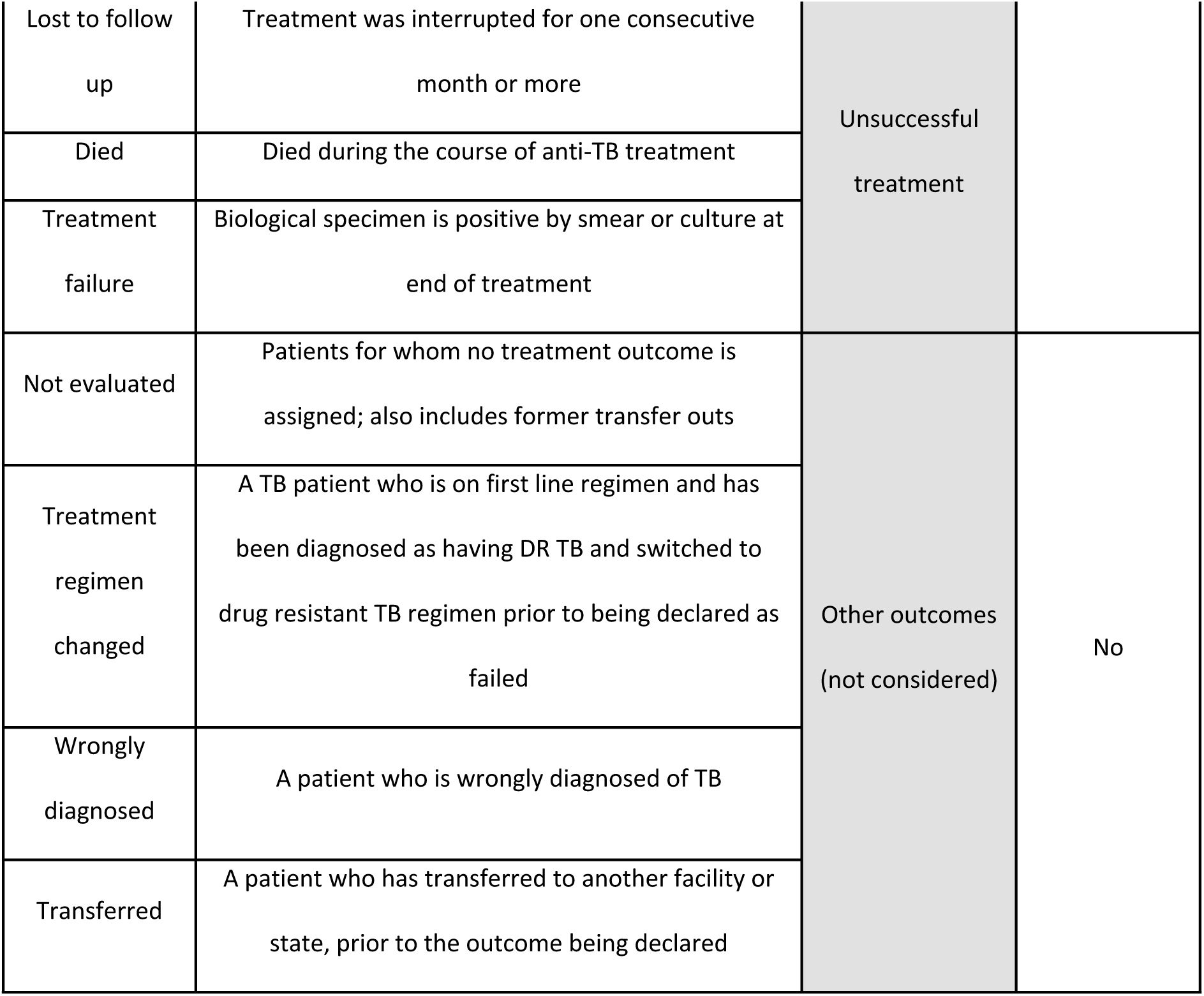
Definitions of treatment outcomes for drug susceptible TB patients [12].

## Appendix 2 Estimating propensity scores

We utilized propensity score modelling [18]–[20] to create a matched dataset comprised of treated patients (CfL) and untreated patients (no CfL), by means of a logistic regression model, including all available potential confounders. The exact model results from the same are given in Table S4, and Table S5 displays the mean propensity scores between patients who actually received CfL or not. The accuracy of the model is 7 77% with a specificity rate of 19%. Figure SF1 shows the distribution of propensity scores visually, segregated by whether or not the patient was on the CfL program.

**Table S2.** Estimating propensity score using logistic regression; N = 989.

| Dependent Variable: CfL |  |
| --- | --- |
| male | 0.955 (0.622, 1.288) |
| Age: 6-15 | 0.612 (-1.036, 2.259) |
| Age: 16-19 | 0.817 (-0.794, 2.428) |
| Age: 20-45 | 0.735 (-0.817, 2.288) |
| Age: 46-65 | 0.338 (-1.248, 1.924) |
| Age: > 65 | 0.240 (-1.466, 1.945) |
| Xpert Testing | 2.539 (2.111, 2.968) |
| Free drugs | 11.113 (10.594, 11.632) |
| Extra Pulmonary status | 1.027 (0.675, 1.379) |
| Facility: St Stephens | 6.075 (5.585, 6.564) |
| Facility: Vinod Karhana | 8.295 (7.795, 8.794) |
| Diag Qtr: 2020 Q1 | 2.708 (2.370, 3.047) |
| Constant | 0.069 (-1.539, 1.678) |
| Observations | 989 |
| Log Likelihood | -440.605 |
| Akaike Inf. Crit. | 907.209 |

Note: a) 95% C.I. based on robust standard errors; b) \*p<0.1; \*\*p<0.05; \*\*\*p<0.01; c) 95% C.I.
displayed alongside results

**Table S3.** Propensity Score estimated by whether or not a patient was on CfL.

| enrolled in CfL | N | Mean score | Median score |
| --- | --- | --- | --- |
| No | 713 | 0.20 | 0.12 |
| Yes | 276 | 0.48 | 0.49 |

**Figure SF1:**
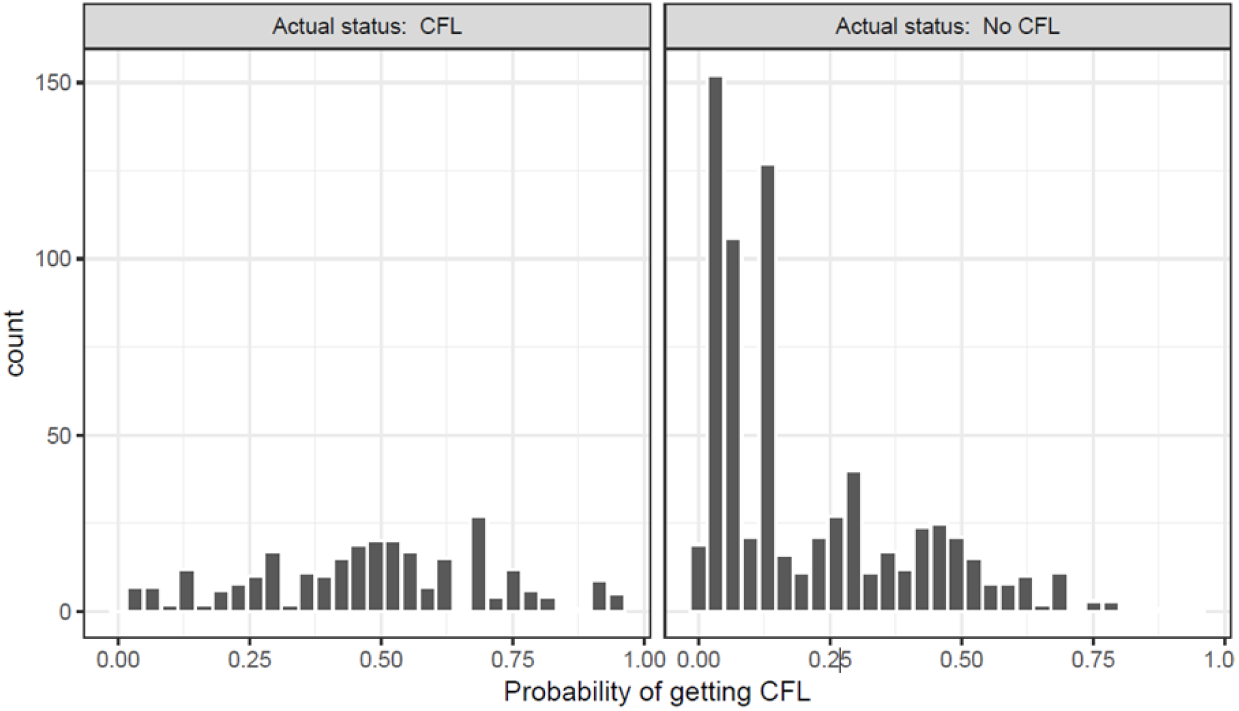
Histograms of the estimated propensity scores by treatment status.

## Appendix 3 Propensity Score Matching

**Table S4.** Summary of balance for the dataset, before and after matching.

|  | Analytical Dataset |  |  |  |  | Matched Dataset |  |  |  |  |
| --- | --- | --- | --- | --- | --- | --- | --- | --- | --- | --- |
|  | Means<br>Treated | Means<br>Control | diff | Std.<br>Mean<br>Diff. | Var.<br>Ratio | Means<br>Treated | Means<br>Control | diff | Std.<br>Mean<br>Diff. | Var.<br>Ratio |
| Diff | 0.48 | 0.20 | 0.28 | 1.25 | 1.37 | 0.44 | 0.44 | 0.00 | 0.00 | 1.00 |
| Age: 0-5 | 0.01 | 0.01 | 0.00 | 0.02 |  | 0.01 | 0.01 | 0.00 | 0.01 |  |
| Age: 6-15 | 0.08 | 0.06 | 0.02 | 0.06 |  | 0.08 | 0.06 | 0.02 | 0.09 |  |
| Age: 16-19 | 0.14 | 0.07 | 0.07 | 0.21 |  | 0.14 | 0.11 | 0.03 | 0.07 |  |
| Age: 20-45 | 0.56 | 0.47 | 0.09 | 0.17 |  | 0.57 | 0.61 | -0.03 | -0.07 |  |
| Age: 46-65 | 0.17 | 0.28 | -0.11 | -0.31 |  | 0.16 | 0.16 | 0.00 | 0.00 |  |
| Age: >65 | 0.04 | 0.11 | -0.07 | -0.33 |  | 0.04 | 0.06 | -0.02 | -0.09 |  |
| Male | 0.54 | 0.57 | -0.03 | -0.07 |  | 0.53 | 0.53 | 0.00 | 0.00 |  |
| Xpert Testing | 0.42 | 0.15 | 0.27 | 0.55 |  | 0.39 | 0.39 | 0.00 | 0.00 |  |
| Free drugs | 0.25 | 0.07 | 0.18 | 0.42 |  | 0.20 | 0.20 | 0.00 | 0.00 |  |
| Facility: sir ganga ram | 0.27 | 0.64 | -0.37 | -0.82 |  | 0.29 | 0.28 | 0.01 | 0.02 |  |
| Facility: st stephens | 0.47 | 0.21 | 0.25 | 0.51 |  | 0.45 | 0.45 | 0.01 | 0.01 |  |
| Facility: vinod karhana | 0.26 | 0.15 | 0.11 | 0.26 |  | 0.26 | 0.27 | -0.01 | -0.03 |  |
| Diag Qtr: 2019 Q4 | 0.36 | 0.52 | -0.16 | -0.34 |  | 0.38 | 0.38 | 0.00 | -0.01 |  |
| Diag Qtr: 2020 Q1 | 0.64 | 0.48 | 0.16 | 0.34 |  | 0.62 | 0.62 | 0.00 | 0.01 |  |
| Extra Pulmonary | 0.50 | 0.57 | -0.08 | -0.15 |  | 0.50 | 0.50 | 0.00 | 0.00 |  |

**Figure SF2:**
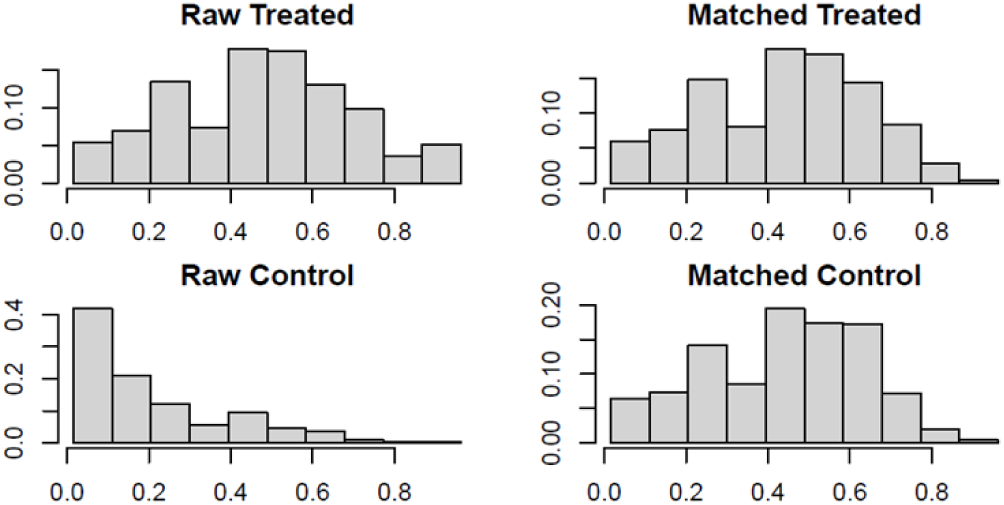
Propensity Scores, before and after the matching, in the treated (CfLTM engagement) and control groups (no CfLTM)

**Figure SF3:**
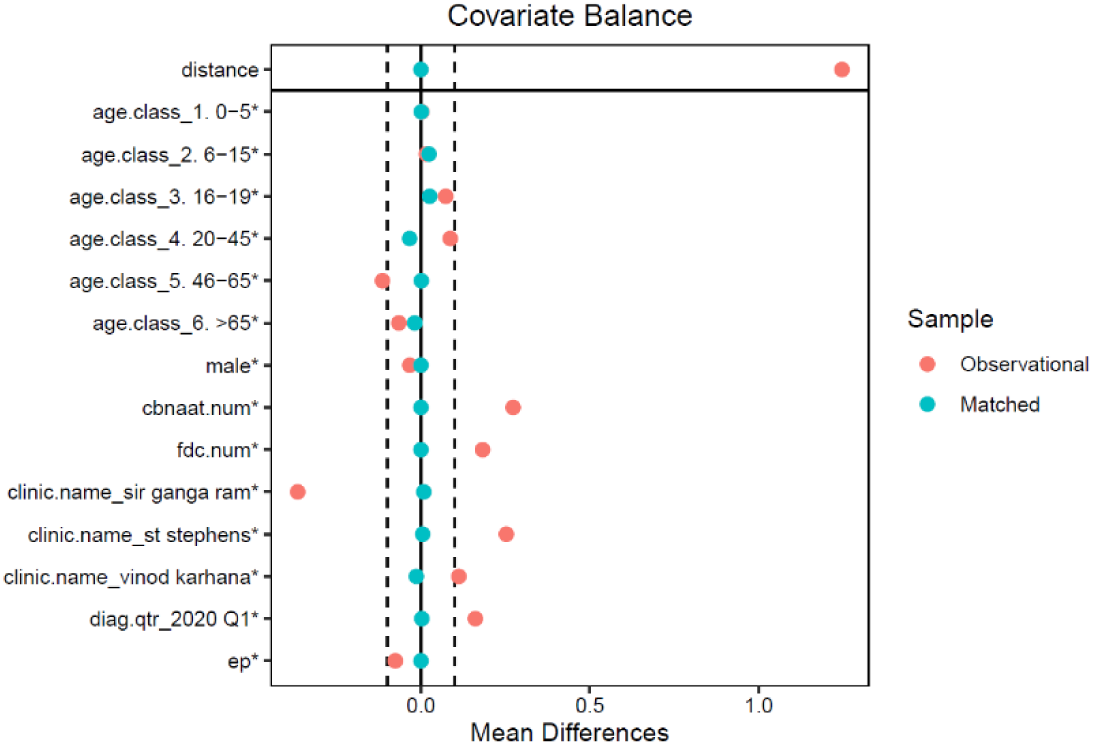
Results from balancing the covariates after the matching procedure. Note: The red dots indicate the differences between standardized means of covariates in the matched and treated groups for the analytical or the unmatched dataset. The green dots indicate the same for the matched or the adjusted dataset

## Appendix 4 Full Model Results; OLS Model; Matched Dataset

**Table S5.** Results from the full OLS model on matched dataset; N = 747.

| Dependent Variable: Follow Ups |  |
| --- | --- |
| CfL | 6.438*** (0.600) |
| Xpert Testing | -0.785 (0.762) |
| Free drugs | -0.972 (0.931) |
| Age: 6-15 | -0.315 (2.291) |
| Age: 16-19 | -0.687 (2.257) |
| Age: 20-45 | -0.328 (2.150) |
| Age: 46-65 | -1.786 (2.182) |
| Age: > 65 | -3.217 (2.303) |
| Male | -0.375 (0.476) |
| Extra Pulmonary | 0.861* (0.507) |
| Facility: st stephens | 2.663*** (0.971) |
| Facility: vinod karhana | 4.679*** (0.977) |
| Diag Qtr: 2020 Q1 | 3.988*** (0.731) |
| Facility: st stephens * Diag Qtr 2020 Q1 | -4.597*** (1.165) |
| Facility: vinod karhana * Diag Qtr 2020 Q1 | -6.818*** (1.239) |
| Constant | 8.185*** (2.251) |
| Observations | 747 |
| R | 0.217 |
| Adjusted R | 0.201 |
| Residual Std. Error | 6.352 (df = 731) |
| F Statistic | 13.526 (df = 15; 731) |

Note: a) 95% C.I. based on robust standard errors; b) \*p<0.1; \*\*p<0.05; \*\*\*p<0.01; c) Model was fitted on the matched dataset; d) We have only 747 observations as 199 patients did not have a recorded value for the number of follow ups conducted

## Appendix 5 Full Model Results; Logistic Model; Matched Dataset

**Table S6.**
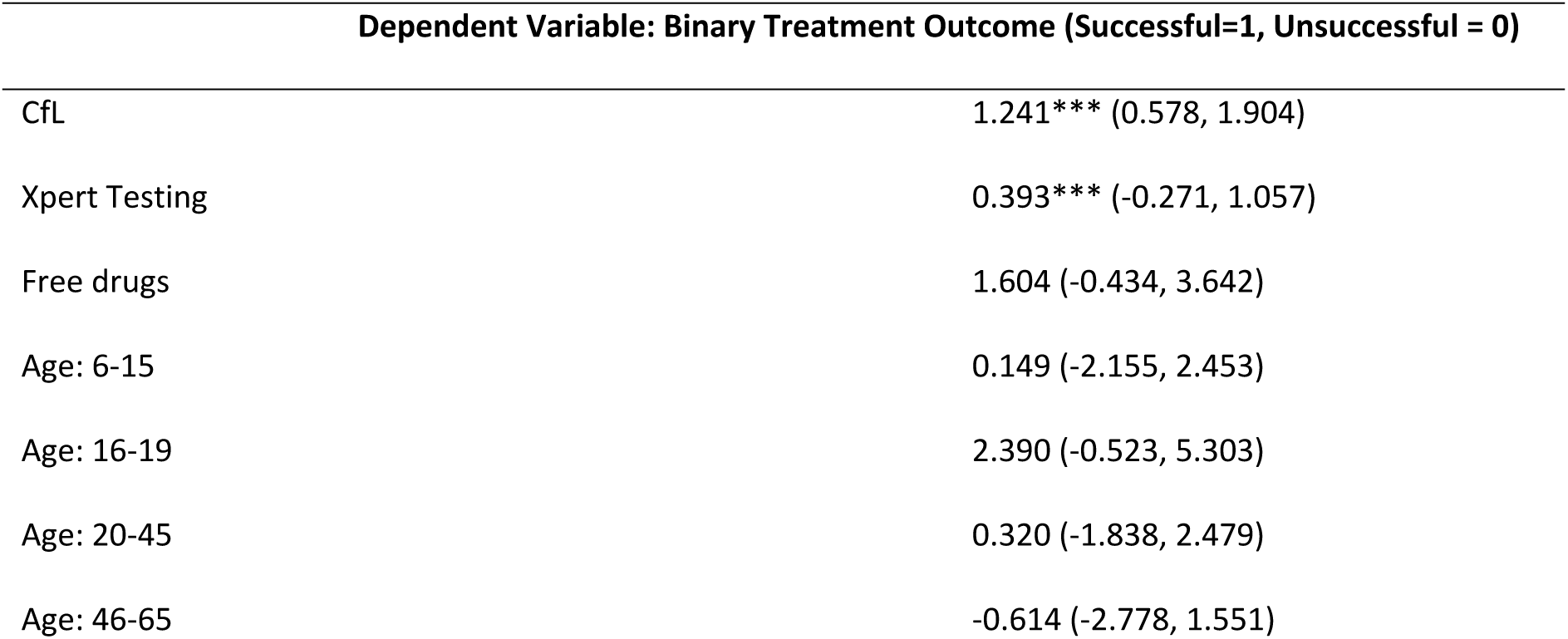
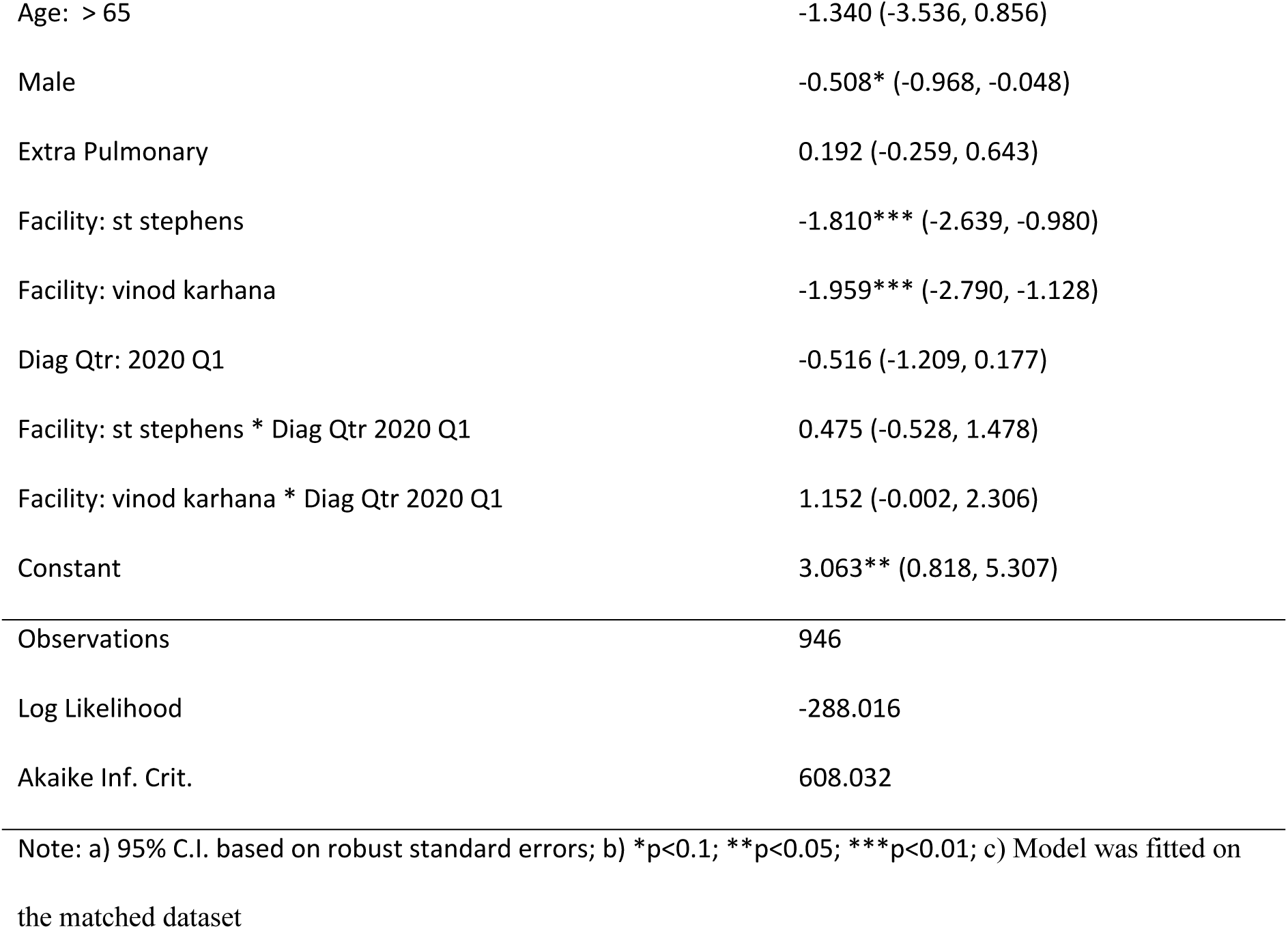
Logistic Regression; Impact of CfL on Treatment Outcomes; N = 946.

## Appendix 6 Sensitivity Analysis; Results from Simple OLS & Logistic Models; Matched and Unmatched (observational) datasets

**Table S9.**
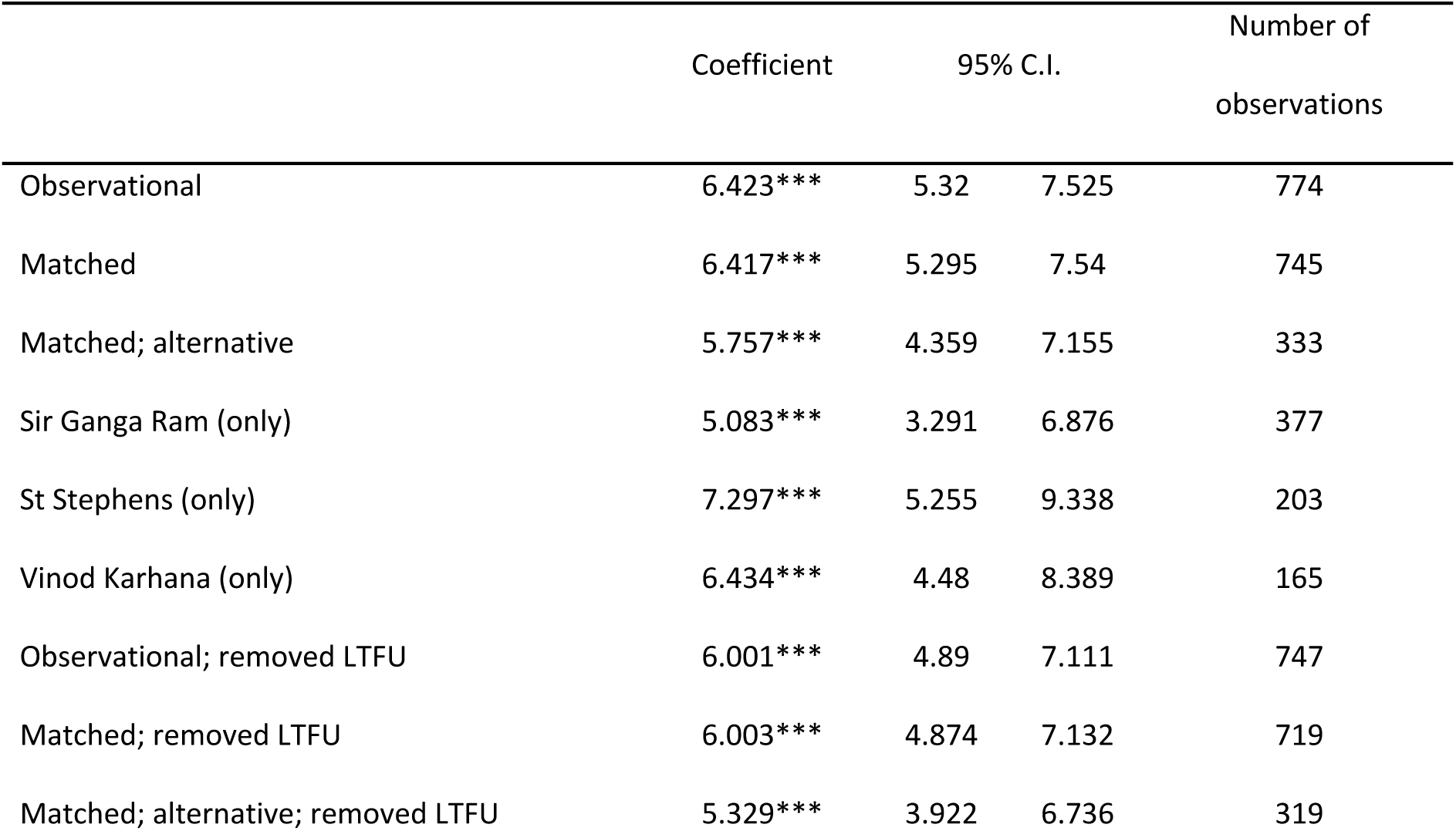
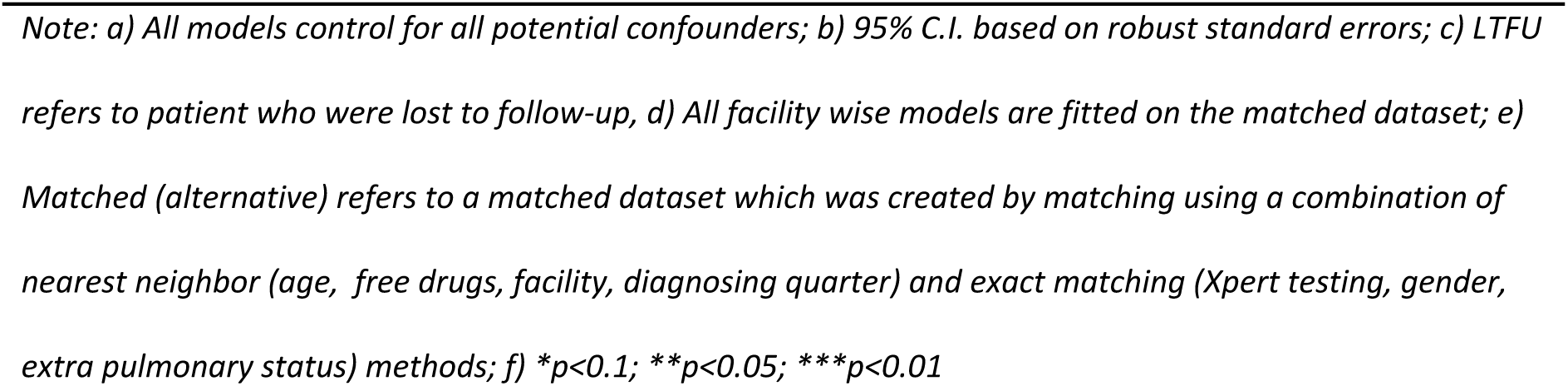
Sensitivity analysis for OLS regression modelling the impact of CfL engagement on number of follow-ups.

|  | Coefficient | 95% C.I. |  | Number of observations |
| --- | --- | --- | --- | --- |
| Observational | 6.423*** | 5.32 | 7.525 | 774 |
| Matched | 6.417*** | 5.295 | 7.54 | 745 |
| Matched; alternative | 5.757*** | 4.359 | 7.155 | 333 |
| Sir Ganga Ram (only) | 5.083*** | 3.291 | 6.876 | 377 |
| St Stephens (only) | 7.297*** | 5.255 | 9.338 | 203 |
| Vinod Karhana (only) | 6.434*** | 4.48 | 8.389 | 165 |
| Observational; removed LTFU | 6.001*** | 4.89 | 7.111 | 747 |
| Matched; removed LTFU | 6.003*** | 4.874 | 7.132 | 719 |
| Matched; alternative; removed LTFU | 5.329*** | 3.922 | 6.736 | 319 |

Note: a) All models control for all potential confounders; b) 95% C.I. based on robust standard errors; c) LTFU refers to patient who were lost to follow-up, d) All facility wise models are fitted on the matched dataset; e) Matched (alternative) refers to a matched dataset which was created by matching using a combination of nearest neighbor (age, free drugs, facility, diagnosing quarter) and exact matching (Xpert testing, gender, extra pulmonary status) methods; f) \* $p<0.1$ ; \*\* $p<0.05$ ; \*\*\* $p<0.01$

**Table S10.** Sensitivity analysis for logistic regression, modelling the impact of the CfL engagement on treatment outcomes.

|  | Odds Ratio (OR) | 95% C.I. (OR) |  | Number of observations |
| --- | --- | --- | --- | --- |
| Observational | 1.255*** | 1.788 | 6.888 | 989 |
| Matched | 1.228*** | 1.701 | 6.857 | 944 |
| Matched; alternative | 1.181*** | 1.563 | 6.791 | 408 |
| Sir Ganga Ram (only) | 0.425 | 0.349 | 6.702 | 519 |
| St Stephens (only) | 1.250*** | 1.405 | 8.677 | 256 |
| Vinod Karhana (only) | 1.562*** | 1.145 | 19.856 | 169 |
| Observational; removed LTFU | 1.319*** | 1.687 | 8.294 | 962 |
| Matched; removed LTFU | 1.319*** | 1.626 | 8.606 | 918 |
| Matched; alternative; removed LTFU | 1.229*** | 1.391 | 8.405 | 394 |

Note: a) All models control for all potential confounders; b) 95% C.I. based on robust standard errors; c) LTFU refers to patient who were lost to follow-up, d) All facility wise models are fitted on the matched dataset; e) Matched (alternative) refers to a matched dataset which was created by matching using a combination of nearest neighbor (age, free drugs, facility, diagnosing quarter) and exact matching (Xpert testing, gender, extra pulmonary status) methods; f) \* $p<0.1$ ; \*\* $p<0.05$ ; \*\*\* $p<0.01$

## Appendix 7 Alternative Matching method

Multiple different matching specifications were run to test for the robustness of the model. We report results for one such alternative matching specification. In this alternative specification, we used the nearest neighbor algorithm to match four covariates, 1) age category, 2) free drug status, 3) facility of diagnosis and 4) diagnosing quarter, and exact matching for 1) proportion of males, 2) proportion of extra pulmonary cases, and 3) proportion of patients diagnosed using Xpert testing. The caliper width used was 0.2. This particular matching resulted in 204 pairs (408 observations), relative to 944 observations obtained by way of full matching. The matching resulted in a more similar set of covariates for the two groups, as observed by the p-values obtained for testing difference between two groups (Table S11).

**Table S11.**
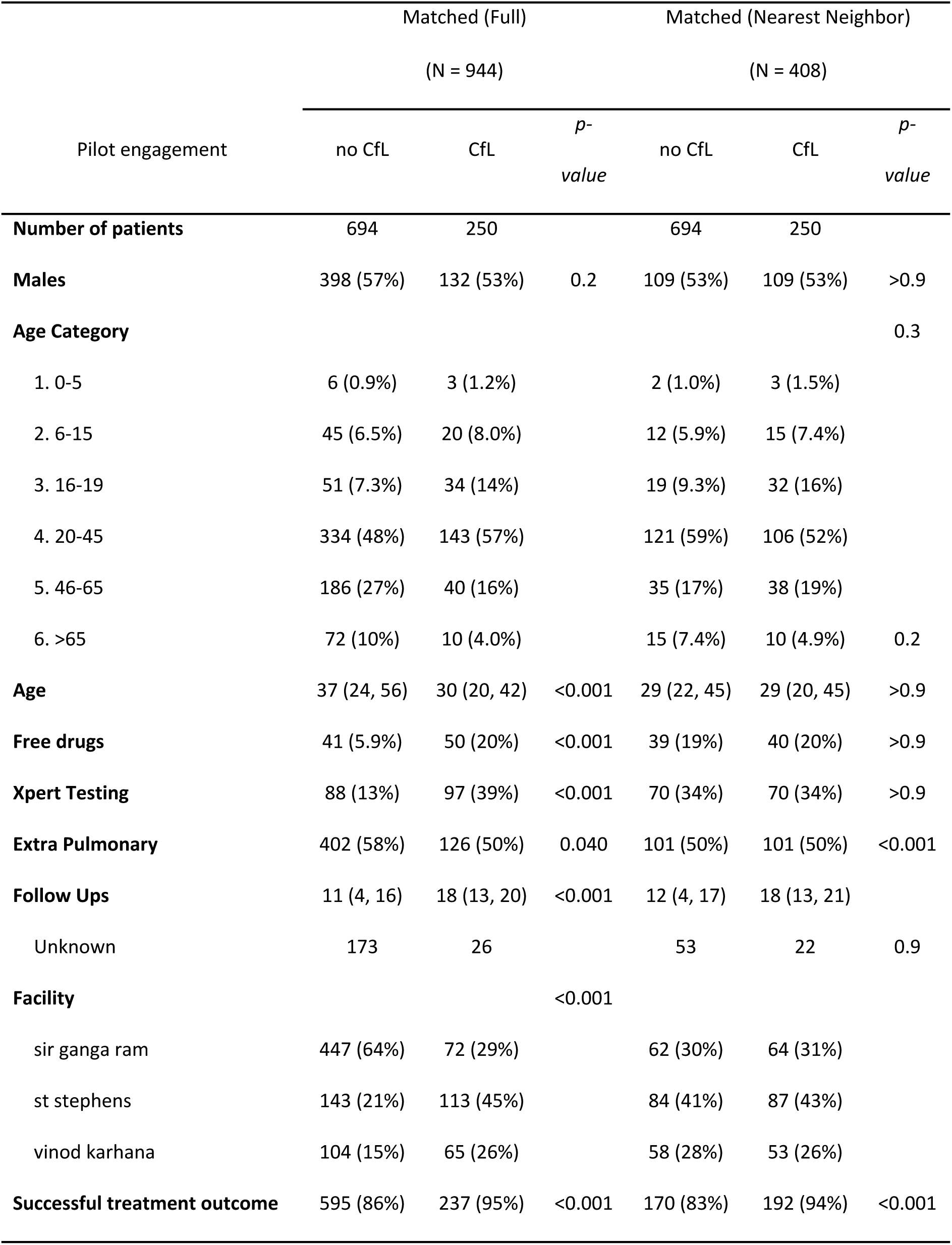
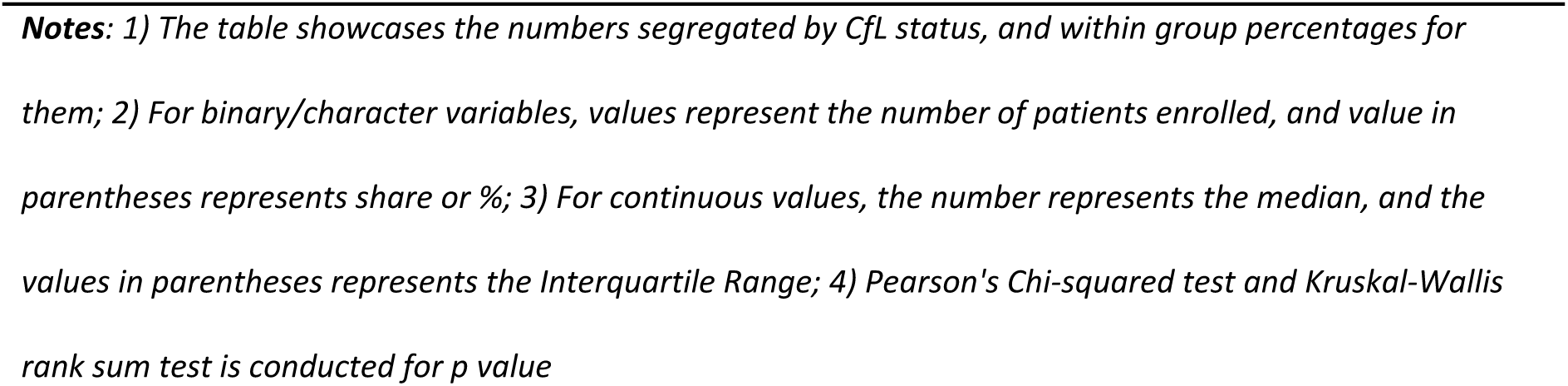
Comparison of descriptive statistics between matched datasets (full matching vs nearest neighbor)

## References

[1] B. K. Ngwatu et al., “The impact of digital health technologies on tuberculosis treatment: a systematic review,” Eur. Respir. J., vol. 51, no. 1, p. 1701596, Jan. 2018, doi: 10.1183/13993003.01596-2017.

[2] A. Ridho et al., “Digital Health Technologies to Improve Medication Adherence and Treatment Outcomes in Patients With Tuberculosis: Systematic Review of Randomized Controlled Trials,” J. Med. Internet Res., vol. 24, no. 2, p. e33062, Feb. 2022, doi: 10.2196/33062.

[3] R. Subbaraman et al., “Digital adherence technologies for the management of tuberculosis therapy: mapping the landscape and research priorities,” BMJ Glob. Health, vol. 3, no. 5, p. e001018, Oct. 2018, doi: 10.1136/bmjgh-2018-001018.

[4] S. Kumar et al., “Mobile Health Technology Evaluation,” Am. J. Prev. Med., vol. 45, no. 2, pp. 228–236, Aug. 2013, doi: 10.1016/j.amepre.2013.03.017.

[5] D. C. Mohr, K. Cheung, S. M. Schueller, C. Hendricks Brown, and N. Duan, “Continuous Evaluation of Evolving Behavioral Intervention Technologies,” Am. J. Prev. Med., vol. 45, no. 4, pp. 517–523, Oct. 2013, doi: 10.1016/j.amepre.2013.06.006.

[6] M. S. Marcolino, J. A. Q. Oliveira, M. D’Agostino, A. L. Ribeiro, M. B. M. Alkmim, and D. Novillo-Ortiz, “The Impact of mHealth Interventions: Systematic Review of Systematic Reviews,” JMIR MHealth UHealth, vol. 6, no. 1, p. e23, Jan. 2018, doi: 10.2196/mhealth.8873.

[7] J. Needamangalam Balaji et al., “A Scoping Review on Accentuating the Pragmatism in the Implication of Mobile Health (mHealth) Technology for Tuberculosis Management in India,” J. Pers. Med., vol. 12, no. 10, p. 1599, Sep. 2022, doi: 10.3390/jpm12101599.

[8] C. M. Denkinger, J. Grenier, A. K. Stratis, A. Akkihal, N. Pant-Pai, and M. Pai, “Mobile health to improve tuberculosis care and control: a call worth making [Review article],” Int. J. Tuberc. Lung Dis., vol. 17, no. 6, pp. 719–727, Jun. 2013, doi: 10.5588/ijtld.12.0638.

[9] W. Tumuhimbise and A. Musiimenta, “A review of mobile health interventions for public private mix in tuberculosis care,” Internet Interv., vol. 25, p. 100417, Sep. 2021, doi: 10.1016/j.invent.2021.100417.

[10] “Joint Effort for Elimination of Tuberculosis.” Accessed: Jun. 28, 2022. [Online]. Available: https://www.projectjeet.in/

[11] “Connect for Life^TM^ (powered by Johnson & Johnson Global Public Health); https://wiki.openmrs.org/display/docs/Connect+for+Life+Distribution.”

[12] Central TB Division, “Treatment Outcomes for drug susceptible TB patients,” in Technical and Operational Guidelines for TB Control in India, 2016, p. 65. [Online]. Available: https://tbcindia.gov.in/showfile.php?lid=3220

[13] C. G. Victora, J.-P. Habicht, and J. Bryce, “Evidence-Based Public Health: Moving Beyond Randomized Trials,” Am J Public Health, vol. 94, pp. 400–5, 2004.

[14] M. S. Ali, D. Prieto-Alhambra, L. C. Lopes, D. Ramos, N. Bispo, and M. Y. Ichihara, “Propensity Score Methods in Health Technology Assessment: Principles, Extended Applications, and Recent Advances,” Front Pharmacol, vol. 10, no. 973, 2019.

[15] A. Cois and R. Ehrlich, “Problem drinking as a risk factor for tuberculosis: a propensity score matched analysis of a national survey,” BMC Public Health, vol. 13, no. 871, 2013.

[16] P. C. Austin, “An Introduction to Propensity Score Methods for Reducing the Effects of Confounding in Observational Studies,” Multivar. Behav Res, vol. 46, pp. 399–424, 2011.

[17] M. Caliendo and S. Kopeinig, “Some Practical Guidance for the Implementation of Propensity Score Matching,” J Econ. Surv., vol. 22, pp. 31–72, 2008.

[18] P. R. Rosenbaum and D. B. Rubin, “The central role of the propensity score in observational studies for causal effects,” Biometrika, vol. 70, no. 1, pp. 41–55, 1983, doi: 10.1093/biomet/70.1.41.

[19] W. C. Winkelmayer and T. Kurth, “Propensity scores: help or hype?,” Nephrol. Dial. Transplant., vol. 19, no. 7, pp. 1671–1673, Jul. 2004, doi: 10.1093/ndt/gfh104.

[20] C. Heinrich, A. Maffioli, and G. Vázquez, “A Primer for Applying Propensity-Score Matching,” Inter-American Development Bank, Office of Strategic Planning and Development Effectiveness (SPD), SPD Working Paper 1005, Aug. 2010. Accessed: Aug. 12, 2022. [Online]. Available: https://econpapers.repec.org/paper/idbspdwps/1005.htm

[21] P. R. Rosenbaum, “A Characterization of Optimal Designs for Observational Studies,” J. R. Stat. Soc. Ser. B Methodol., vol. 53, no. 3, pp. 597–610, 1991, doi: http://www.jstor.org/stable/2345589.

[22] B. B. Hansen, “Full Matching in an Observational Study of Coaching for the SAT,” J. Am. Stat. Assoc., vol. 99, no. 467, pp. 609–618, Sep. 2004, doi: 10.1198/016214504000000647.

[23] K. Ming and P. R. Rosenbaum, “Substantial Gains in Bias Reduction from Matching with a Variable Number of Controls,” Biometrics, vol. 56, no. 1, pp. 118–124, Mar. 2000, doi: 10.1111/j.0006-341X.2000.00118.x.

[24] P. C. Austin, “Optimal caliper widths for propensity-score matching when estimating differences in means and differences in proportions in observational studies,” Pharm Stat, vol. 10, pp. 150–61, 2011.

[25] H. O. Balli and B. E. Sørensen, “Interaction effects in econometrics,” Empir. Econ., vol. 45, no. 1, pp. 583–603, Aug. 2013, doi: 10.1007/s00181-012-0604-2.

[26] Q.-Y. Zhao, J.-C. Luo, Y. Su, Y.-J. Zhang, G.-W. Tu, and Z. Luo, “Propensity score matching with R: conventional methods and new features,” Ann. Transl. Med., vol. 9, pp. 812–812, 2021.

[27] D. Ho, K. Imai, G. King, and S. E. A. MatchIt, “Nonparametric Preprocessing for Parametric Causal Inference,” J. Stat. Softw., vol. 42, pp. 1–28, 2011.

[28] S. Selvaraj, K. Karan, S. Srivastava, N. Bhan, and I. Mukhopadhyay, “India Health System Review.” World Health Organization, Regional Office for South-East Asia, 2022. [Online]. Available: https://apo.who.int/publications/i/item/india-health-system-review

[29] R. Danasekaran, T. Raja, and B. Kumar M, “mHealth: A Newer Perspective in Healthcare through Mobile Technology,” J. Compr. Health, vol. 7, no. 2, pp. 67–68, Dec. 2019, doi: 10.53553/JCH.v07i02.012.

[30] A. Musiimenta et al., “Mobile Health Technologies May Be Acceptable Tools for Providing Social Support to Tuberculosis Patients in Rural Uganda: A Parallel Mixed-Method Study,” Tuberc. Res. Treat., vol. 2020, pp. 1–8, Jan. 2020, doi: 10.1155/2020/7401045.

[31] I. Margineanu et al., “Patients and Medical Staff Attitudes Toward the Future Inclusion of eHealth in Tuberculosis Management: Perspectives From Six Countries Evaluated using a Qualitative Framework,” JMIR MHealth UHealth, vol. 8, no. 11, p. e18156, Nov. 2020, doi: 10.2196/18156.

[32] A. I. Latif, E. L. Sjattar, and K. A. Erika, “Models and benefits of mobile health application to support patient with tuberculosis: A literature review,” Enferm. Clínica, vol. 30, pp. 163–167, Mar. 2020, doi: 10.1016/j.enfcli.2019.07.069.

[33] N. Arinaminpathy et al., “Modelling the potential impact of adherence technologies on tuberculosis in India,” Int. J. Tuberc. Lung Dis., vol. 24, no. 5, pp. 526–533, May 2020, doi: 10.5588/ijtld.19.0472.

[34] S. B. Nagaraja et al., “‘Kill-TB’ Drug Reminder Mobile Application for Tuberculosis Patients at Bengaluru, India: Effectiveness and Challenges,” J. Tuberc. Res., vol. 08, no. 01, pp. 1–10, 2020, doi: 10.4236/jtr.2020.81001.

[35] S. Oberoi, V. K. Gupta, N. Chaudhary, and A. Singh, “99 DOTS Mini Review,” ICJMR, 2016, [Online]. Available: https://www.researchgate.net/publication/315542622_99_DOTS_MINI-REVIEW

[36] World Health Organization, Handbook for the use of digital technologies to support tuberculosis medication adherence. Geneva: World Health Organization, 2017. Accessed: Feb. 01, 2023. [Online]. Available: https://apps.who.int/iris/handle/10665/259832

[37] X. Liu et al., “Usability of a Medication Event Reminder Monitor System (MERM) by Providers and Patients to Improve Adherence in the Management of Tuberculosis,” Int. J. Environ. Res. Public. Health, vol. 14, no. 10, p. 1115, Sep. 2017, doi: 10.3390/ijerph14101115.

[38] S. Saha et al., “Tuberculosis Monitoring Encouragement Adherence Drive (TMEAD): Toward improving the adherence of the patients with drug-sensitive tuberculosis in Nashik, Maharashtra,” Front. Public Health, vol. 10, p. 1021427, Dec. 2022, doi: 10.3389/fpubh.2022.1021427.

[39] B. E. Thomas et al., “Acceptability of the Medication Event Reminder Monitor for Promoting Adherence to Multidrug-Resistant Tuberculosis Therapy in Two Indian Cities: Qualitative Study of Patients and Health Care Providers,” J. Med. Internet Res., vol. 23, no. 6, p. e23294, Jun. 2021, doi: 10.2196/23294.

[40] T. Manyazewal, Y. Woldeamanuel, D. P. Holland, A. Fekadu, and V. C. Marconi, “Effectiveness of a digital medication event reminder and monitor device for patients with tuberculosis (SELFTB): a multicenter randomized controlled trial,” BMC Med., vol. 20, no. 1, p. 310, Sep. 2022, doi: 10.1186/s12916-022-02521-y.

[41] D. Drabarek, N. T. Anh, N. V. Nhung, N. B. Hoa, G. J. Fox, and S. Bernays, “Implementation of Medication Event Reminder Monitors among patients diagnosed with drug susceptible tuberculosis in rural Viet Nam: A qualitative study,” PLOS ONE, vol. 14, no. 7, p. e0219891, Jul. 2019, doi: 10.1371/journal.pone.0219891.

[42] J. Acosta, P. Flores, M. Alarcón, M. Grande-Ortiz, L. Moreno-Exebio, and Z. M. Puyen, “A randomised controlled trial to evaluate a medication monitoring system for TB treatment,” Int. J. Tuberc. Lung Dis., vol. 26, no. 1, pp. 44–49, Jan. 2022, doi: 10.5588/ijtld.21.0373.

[43] A. Cross et al., “99DOTS: a low-cost approach to monitoring and improving medication adherence,” in Proceedings of the Tenth International Conference on Information and Communication Technologies and Development, Ahmedabad India: ACM, Jan. 2019, pp. 1–12. doi: 10.1145/3287098.3287102.

[44] D. Thakkar, K. Piparva, and S. Lakkad, “A pilot project: 99DOTS information communication technology-based approach for tuberculosis treatment in Rajkot district,” Lung India, vol. 36, no. 2, p. 108, 2019, doi: 10.4103/lungindia.lungindia_86_18.

[45] N. Alipanah et al., “Adherence interventions and outcomes of tuberculosis treatment: A systematic review and meta-analysis of trials and observational studies,” PLOS Med., vol. 15, no. 7, p. e1002595, Jul. 2018, doi: 10.1371/journal.pmed.1002595.

[46] X. Liu et al., “Effectiveness of Electronic Reminders to Improve Medication Adherence in Tuberculosis Patients: A Cluster-Randomised Trial,” PLOS Med., vol. 12, no. 9, p. e1001876, Sep. 2015, doi: 10.1371/journal.pmed.1001876.

[47] P. Thekkur et al., “Outcomes and implementation challenges of using daily treatment regimens with an innovative adherence support tool among HIV-infected tuberculosis patients in Karnataka, India: a mixed-methods study,” Glob. Health Action, vol. 12, no. 1, p. 1568826, Jan. 2019, doi: 10.1080/16549716.2019.1568826.

[48] N. K. Jose, C. Vaz, P. R. Chai, and R. Rodrigues, “The Acceptability of Adherence Support via Mobile Phones for Antituberculosis Treatment in South India: Exploratory Study,” JMIR Form. Res., vol. 6, no. 5, p. e37124, May 2022, doi: 10.2196/37124.

[49] R. Sodhi, M. J. Penkunas, and A. Pal, “Free drug provision for tuberculosis increases patient follow-ups and successful treatment outcomes in the Indian private sector: A quasi experimental study using propensity score matching,” In Review, preprint, Jan. 2023. doi: 10.21203/rs.3.rs-2448126/v1.

[50] Z. Shubber et al., “Patient-Reported Barriers to Adherence to Antiretroviral Therapy: A Systematic Review and Meta-Analysis,” PLOS Med., vol. 13, no. 11, p. e1002183, Nov. 2016, doi: 10.1371/journal.pmed.1002183.

[51] S. A. Munro, S. A. Lewin, H. J. Smith, M. E. Engel, A. Fretheim, and J. Volmink, “Patient Adherence to Tuberculosis Treatment: A Systematic Review of Qualitative Research,” PLoS Med., vol. 4, no. 7, p. e238, Jul. 2007, doi: 10.1371/journal.pmed.0040238.

[52] S. J. Iribarren, R. Schnall, P. W. Stone, and A. Carballo-Diéguez, “Smartphone Applications to Support Tuberculosis Prevention and Treatment: Review and Evaluation,” JMIR MHealth UHealth, vol. 4, no. 2, p. e25, May 2016, doi: 10.2196/mhealth.5022.

[53] D. J Carter et al., “The impact of a cash transfer programme on tuberculosis treatment success rate: a quasi-experimental study in Brazil,” BMJ Glob. Health, vol. 4, no. 1, p. e001029, Jan. 2019, doi: 10.1136/bmjgh-2018-001029.

[54] D. L. Sackett, Ed., Evidence-based medicine: how to practice and teach EBM, 2nd ed., Reprinted. Edinburgh: Churchill Livingstone, 2001.

